# Strategic Treatment Optimization for HCV (STOPHCV1): a randomised controlled trial of ultrashort duration therapy for chronic hepatitis C

**DOI:** 10.1101/2021.01.22.21250208

**Authors:** Graham S. Cooke, Sarah Pett, Leanne McCabe, Chris Jones, Richard Gilson, Sumita Verma, Stephen D Ryder, Jane D Collier, Stephen T. Barclay, Aftab Ala, Sanjay Bhagani, Mark Nelson, Chinlye Ch’Ng, Ben Stone, Martin Wiselka, Daniel Forton, Stuart McPherson, Rachel Halford, Dung Nguyen, David Smith, Azim Ansari, Emily Dennis, Fleur Hudson, Eleanor J Barnes, Ann Sarah Walker, on behalf of the STOP-HCV trial team

## Abstract

**Background:** The WHO has identified the need for a better understanding of which patients can be cured with ultrashort course hepatitis C (HCV) therapy

**Methods:** 202 individuals with chronic HCV were randomised to fixed-duration shortened therapy (8 weeks) vs variable-duration ultrashort strategies (VUS1/2). Participants not cured following first-line treatment were retreated with 12 weeks’ sofosbuvir/ledipasvir/ribavirin. Primary outcome was sustained virological response 12 weeks (SVR12) after first-line treatment and retreatment. Participants were factorially randomised to receive ribavirin with first-line treatment.

**Results:** All evaluable participants achieved SVR12 overall (197/197, 100%[95%CI 98-100]) demonstrating non-inferiority between fixed-duration and variable-duration strategies (difference 0% [95%CI −3.8%,+3.7%], 4% pre-specified non-inferiority margin). First-line SVR12 was 91%[86%-97%] (92/101) for fixed-duration vs 48%[39%-57%] (47/98) for variable-duration, but was significantly higher for VUS2 (72% [56%-87%] (23/32)) than VUS1 (36% [25%-48%] (24/66)). Overall first-line SVR12 was 72%[65%-78%] (70/101) without ribavirin and 68%[61%-76%] (69/98) with ribavirin (p=0.48). At treatment failure, the emergence of viral resistance was lower with ribavirin (12% [2%-30%] (3/26)) than without (38% [21%-58%] (11/29), p=0.01).

**Conclusions:** Unsuccessful first-line short-course therapy did not compromise retreatment with sofosbuvir/ledipasvir/ribavirin (100% SVR12). SVR12 rates were significantly increased when ultrashort treatment varied between 4-7 weeks rather than 4-6 weeks. Ribavirin significantly reduced resistance emergence in those failing first-line therapy.

**Registration:** ISRCTN 37915093.

**Funding:** National Institutes of Health Research.

## Introduction

The recent and rapid development of treatment for hepatitis C virus (HCV) has enabled an ambitious strategy for the elimination of viral hepatitis as a global public health threat by 2030, with the target of treating 80% of those chronically infected with HCV.^1^ Although licensed durations of 8-12 weeks’ therapy with directly acting antivirals (DAAs) are significantly shorter, more tolerable and more effective than previous interferon-based therapies,^2^ there are patients who still find it challenging to complete a full treatment course. Such patients will become an increasingly important part of clinical practice as treatment coverage expands to reach marginalised groups and WHO treatment guidelines highlight the need to understand the factors that could be used to select patients for successful short course treatment^3^.

Shorter treatment courses of licensed therapies are likely to improve adherence, including in those with active illicit drug use.^4,5^ In acute or recent HCV infection, shortened courses of licensed therapy may have sufficiently high efficacy to be recommended routinely.^6^ However, in chronic infection there is limited data to select patients able to achieve high cure rates with short duration therapy. Unselected short duration treatment was able to achieve cure rates of 20-40% with 4 weeks and 57-95% with 6 weeks treatment in small Phase II studies^7 8,9^, but few of these combinations or durations were subsequently licensed for use. For licensed therapies, baseline viral load (<6,000,000 IU/mL)^10^ and subgenotype^11^ have been recommended to shorten therapy from 12 to 8 weeks, but there are no validated criteria to recommend less than 8 weeks therapy in chronic infection.

For a clinician either deciding to start treatment in a patient considered at high risk of not completing therapy, there is a potential concern that emerging resistance with virological failure may compromise future treatment options. However, it is also possible that with shorter courses of treatment, less resistance may emerge. In vitro evidence suggests the addition of ribavirin, a generically available guanosine analogue, may improve rates of virological cure with shorter treatment courses ^12^ and could reduce the emergence of resistance in those failing treatment when added to short-course therapy.^13^ However, these hypotheses have not been tested in a randomised trial.

We performed a strategic post-licensing randomised controlled trial in HCV-infected participants with mild liver disease to evaluate strategies for short-course treatment, the impact of treatment failure on retreatment and the role of adjunctive ribavirin in short-course therapy.

## METHODS

### Design

We conducted a multi-centre, randomised, open-label, factorial, parallel group non-inferiority trial of adults with mild chronic HCV in 14 UK centres (**Figure S1**). The trial was approved by the Cambridgeshire South Research Ethics Committee (15/EE/0435).

### Participants

Adults (≥18 years) infected with HCV genotype 1a/1b/4 for ≥6 months, with consistently detectable viremia 6 months before randomisation, no evidence of significant liver fibrosis (Fibroscan score ≤7.1kPa equivalent to F0-F1^14^), body mass index (BMI) ≥18kg/m^2^, HCV viral load (VL) detectable but <10 million IU/ml at screening, no previous DAA exposure for current infection (previous pegylated-interferon/ribavirin allowed). Individuals co-infected with HIV were eligible if HIV VL had been <50 copies/ml for >24 weeks on anti-HIV drugs. Full inclusion/exclusion criteria are provided in the **Supplementary Methods**. Participants gave written informed consent.

### Randomisation

Participants were randomised 1:1 to variable ultrashort-course treatment strategy (VUS) vs fixed 56 days of first-line treatment. Individuals were also randomised 1:1 using a factorial design to adjunctive ribavirin vs no ribavirin with first-line therapy. Randomisation determined duration of first-line therapy rather than choice of DAAs which was pre-specified by the investigator before randomisation based on local availability from (i) [genotype 1a/1b] co-formulated ombitasvir/paritaprevir/ritonavir once daily plus separate dasabuvir once-daily (total 25mg/150mg/100 mg plus 500mg respectively) (ii) [genotype 4] ombitasvir/paritaprevir/ritonavir 25/150/100mg once-daily (iii) [genotype 1a/1b/4] glecaprevir/pibrentasvir 300/120mg once-daily (only available after 1 November 2017). Ribavirin dosing was weight-based twice-daily (<75kg 1000mg/day, ≥75kg 1200mg/day). Full details within **Supplementary Methods**.

### Intervention and procedures

For participants allocated to VUS, the duration of first-line therapy varied between 28 and 42 (mean 32; before 1 April 2017) or 49 (mean 39; after 1 April 2017) days determined by the baseline screening VL using a continuous scale (**Tables S1, S2, Figure S2)**. Blinding was not used because the primary end-point was an objective measure of viraemia blinded to clinical data measured in routine laboratories without knowledge of randomisation.

All patients failing treatment were retreated as soon as practicable with 12 weeks of sofosbuvir 400mg/ledipasvir 90mg once-daily and weight-based ribavirin twice-daily (dosing as above). Treatment failure was defined as (i) two consecutive measurements of HCV VL above the lower limit of quantification (LLOQ) (taken at least one week apart) after two consecutive visits with HCV VL <LLOQ at any time, with the latter confirmatory measurement also being >2000 IU/mL, or (ii) two consecutive measurements of HCV VL (taken at least one week apart) that were >1log_10_ increase above the nadir on treatment and >2000 IU/mL at any time.

### Outcomes

The primary outcome was Sustained Virological Response 12 (SVR12, plasma HCV VL <LLOQ without prior failure 12 weeks after the end of the combined first and any re-treatment phases). For ribavirin comparison the primary outcome was SVR12 after first-line treatment only. Secondary outcomes were SVR12 after first-line treatment (where not the primary outcome), SVR12 after the end of the combined first and any re-treatment phases (where not the primary outcome), SVR24 after the end of the combined first and any re-treatment phases, SVR24 after first-line treatment only, lack of initial virological response, viral load rebound after becoming undetectable, serious adverse events, grade 3/4 adverse events, grade 3/4 adverse events judged definitely/probably related to interventions, treatment-modifying adverse events (any grade), grade 3/4 anaemia and emergence of resistance-associated HCV variants. Adverse events were graded following the Division of AIDS grading tables.^15^ HCV genome sequences were generated using next generation sequencing with probe enrichment in a single laboratory as previously described.^16^

### Statistical analysis

*A priori* power calculation assumed 88% would achieve SVR12 on first-line fixed-duration, and that SVR12 would be 85% on retreatment (significantly lower than the actual retreatment success rate, below), leading to an overall cure rate of 98% (first-line plus retreatment) in the control group. Assuming 98% cure rate in the fixed-duration group, 80% power, one-sided alpha 0.025, and a 5% loss to follow-up, 408 participants were needed to demonstrate non-inferiority with a 4% margin. The choice of the 4% margin was based on clinical judgement and to ensure that overall cure rates in the variable-duration group would be well over 90% if non-inferiority was demonstrated. Interim data were reviewed by an independent Data Monitoring Committee (DMC) (4 biannual meetings). The protocol stipulated that the DMC could alter the first-line treatment strategy if there was strong evidence the SVR12 rate for VUS1 was less than 65%. After the DMC meeting in April 2017, the DAA duration strategy was changed from 4-6 weeks (VUS1) to 4-7 weeks (VUS2) (**Table S1**). All participants randomised from the 1st April 2017 were treated under VUS2. The trial closed in August 2018 when no further recruitment was possible. By this time, the great majority of patients with viraemia were unable to engage with treatment *per se*, and not suitable for inclusion in this study.

Randomised groups were compared following the principle of intention-to-treat (including all follow-up regardless of changes to treatment) using binomial regression (risk difference scale) to estimate risk differences for binary outcomes, Kaplan-Meier and Cox regression for time to first-line failure (with competing risks methods for its components, primary failure and VL rebound), and generalised estimating equations with independent working correlation for global tests of repeated measures (adjusted for baseline for continuous measures). Analyses used Stata v15.1 (additional details in the **Supplementary Methods)**. No adjustment was made for multiple testing. Analyses used Stata v15.1. The trial was registered at ISRCTN, number 37915093.

## RESULTS

Between 18 March 2016 and 28 August 2018, 204 participants from 14 United Kingdom centres were randomised (**Figure 1**). Two participants were randomised in error and excluded, leaving 202 (102 fixed-duration, 100 variable-duration; 100 ribavirin, 102 no-ribavirin) participants in the analyses.

**Figure 1.**
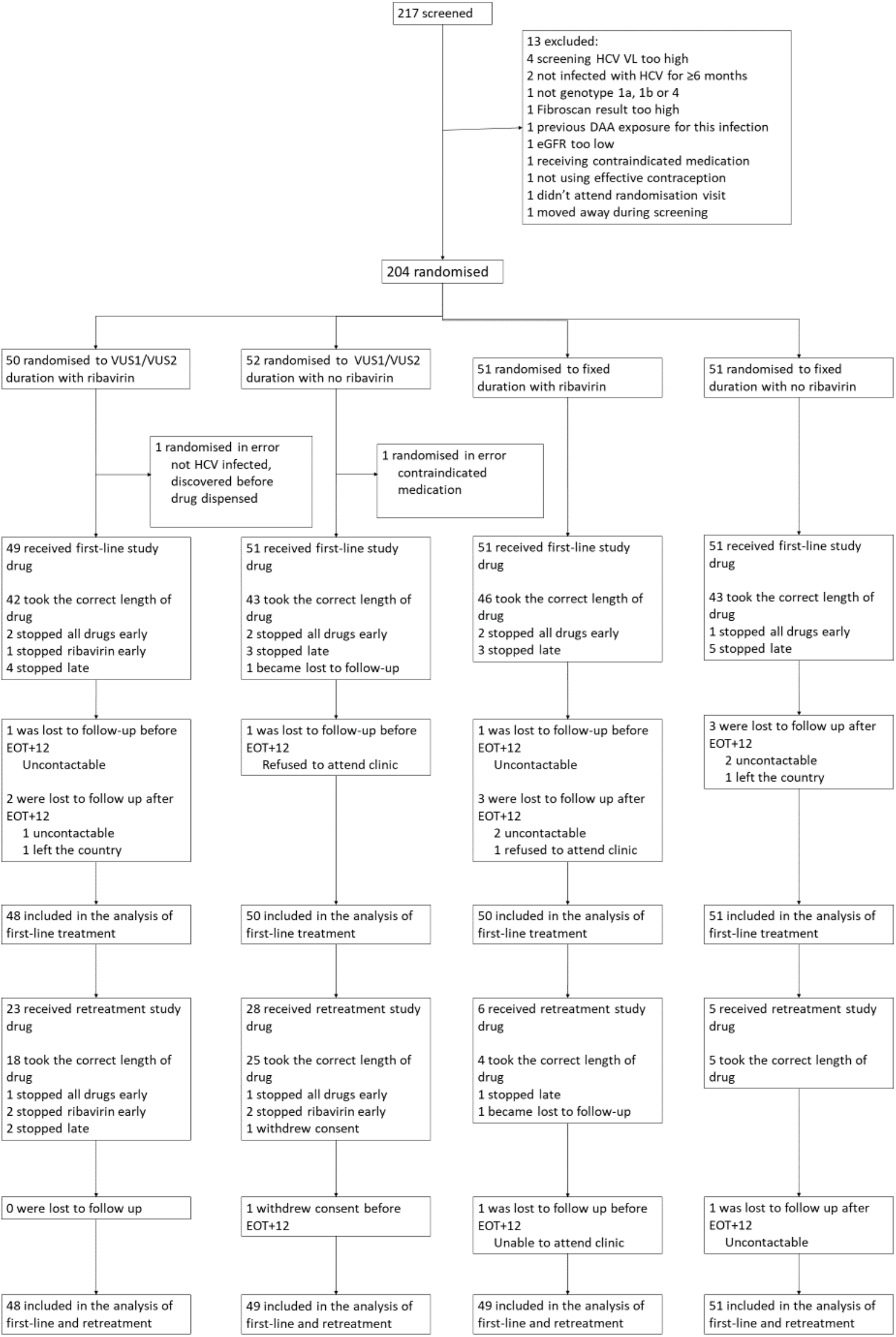
Trial profile. Note: EOT+12: 12 weeks after the end of treatment. Some patients lost to follow up are included in the analysis based on local VLs from medical notes.

Baseline characteristics were broadly balanced between randomised groups (**Tables 1, S3**) and across strategies (before vs after 1 April 2017) (**Table S4**). Median screening and enrolment HCV VL were 711423 and 741946 IU/mL in samples taken a median (IQR) 19 (13,33) days apart. Whilst the median (IQR) difference was 0.01 (−0.19,+0.21) log_10_ IU/mL, absolute differences were greater (**Figure S3**), leading to 57 (28%) participants being excluded from the per-protocol analysis because they would have received a difference of ≥3 days of DAAs had this been determined by the enrolment rather than the screening VL (31 (23%) VUS1 vs 26 (39%) VUS2, because the second strategy received more drug overall, **Figure S2**).

**Table 1.** Characteristics at randomisation.

|  | Total (N=202) | VUS duration<br>(N=100) | Fixed duration<br>(N=102) | Ribavirin<br>(N=100) | No ribavirin<br>(N=102) |
| --- | --- | --- | --- | --- | --- |
| Randomised under first protocol (VUS1) | 136 (67%) | 68 (68%) | 68 (67%) | 68 (68%) | 68 (67%) |
| Age (years) | 45.5 (37.5, 53.0) | 45.2 (38.8, 51.6) | 46.3 (36.6, 54.1) | 46.1 (36.7, 52.4) | 44.8 (37.7, 54.1) |
| Female at birth | 62 (31%) | 28 (28%) | 34 (33%) | 34 (34%) | 28 (27%) |
| BMI (kg/m<sup>2</sup>) | 24.9 (22.2, 27.2) | 24.9 (22.6, 26.7) | 24.9 (21.8, 27.7) | 23.7 (21.7, 26.5) | 25.8 (23.3, 27.6) |
| White ethnicity | 176 (87%) | 89 (89%) | 87 (85%) | 89 (89%) | 87 (85%) |
| Screening HCV viral load (IU/ml) | 711423<br>(218776,1995262) | 790664 (214388,<br>1917731) | 687916 (220000,<br>2381846) | 700272 (169717,<br>2071064) | 750523 (275000,<br>1949844) |
| Enrolment HCV viral load (IU/ml) n=199 | 741946<br>(249097,1872136) | 801000 (251188,<br>1500000) | (614047 (248000,<br>2238721) | 657858 (178842,<br>1500000) | 801000 (385595,<br>2200000) |
| HCV genotype/subgenotype: 1a | 166 (82%) | 82 (82%) | 84 (82%) | 84 (84%) | 82 (80%) |
| 1b | 34 (17%) | 17 (17%) | 17 (17%) | 16 (16%) | 18 (18%) |
| 4 | 2 (1%) | 1 (1%) | 1 (1%) | 0 | 2 (2%) |
| HIV coinfectd | 68 (34%) | 32 (32%) | 36 (35%) | 35 (35%) | 33 (32%) |
| Fibroscan result (kPa) | 4.9 (4.2, 5.8) | 5.0 (4.3, 5.9) | 4.8 (4.1, 5.5) | 4.8 (4.4, 5.8) | 4.9 (4.1, 5.9) |
| Haemoglobin (g/dl) | 14.7 (14.0, 15.6) | 14.8 (14.1, 15.6) | 14.7 (13.8, 15.6) | 14.7 (13.8, 15.6) | 14.8 (14.0, 15.7) |
| ALT (IU/ml) | 52 (34, 87) | 50 (34, 90) | 54 (34, 87) | 51 (35, 89) | 54 (31, 87) |
| IL28b genotype*: CC | 60 (30%) | 32 (32%) | 28 (27%) | 29 (29%) | 31 (30%) |
| CT | 106 (52%) | 51 (51%) | 55 (54%) | 56 (56%) | 50 (49%) |
| TT | 27 (13%) | 14 (14%) | 13 (13%) | 11 (11%) | 16 (16%) |
| No result | 9 (4%) | 3 (3%) | 6 (6%) | 4 (4%) | 5 (5%) |
| Previously unsuccessfully treated with interferon<br>and/or ribavirin | 24 (12%) | 12 (12%) | 12 (12%) | 11 (11%) | 13 (13%) |
| Current/recent alcoholism/alcohol abuse | 13 (6%) | 5 (5%) | 8 (8%) | 7 (7%) | 6 (6%) |
| Current/recent illicit substance abuse | 64 (32%) | 31 (31%) | 33 (32%) | 28 (28%) | 26 (25%) |
| Treated with paritaprevir\ombitasvir\dasabuvir | 198 (98%) | 98 (98%) | 100 (98%) | 100 (100%) | 98 (96%) |
| Treated with paritaprevir\ombitasvir | 2 (1%) | 1 (1%) | 1 (1%) | 0 | 2 (2%) |
| Treated with glecaprevir\pibrentasvir | 2 (1%) | 1 (1%) | 1 (1%) | 0 | 2 (2%) |

\* Result from whole genome sequencing or from Epistem point of care test if genotyping result not available.
Note: showing n (%) for categorical factors, or median (IQR) for continuous factors. Missing data indicated by denominators in the row label. As an indicator of imbalance, $P > 0.05$ for all comparisons of baseline characteristics between groups other than BMI ( $p < 0.001$ ) between ribavirin and no ribavirin groups.

### Follow-up and treatment received

Each pre- or post first-line EOT visit was missed by no more than 8 (4%) participants. One participant was lost-to-follow-up at day-28 on first-line and three (all randomised to fixed-duration, two with ribavirin) stopped first-line treatment >1 week early. All other participants completed first-line treatment(**Figure 2**). Adverse events caused one participant to stop both first-line DAAs and ribavirin 3 days early (grade 3 mouth sores), and one ribavirin only 2 days early (grade 3 anaemia). Self-reported non-adherence to DAAs and/or ribavirin varied from 2-14% across first-line visits (**Figure S4(a)(b)**), with 55 (28%) reported missing doses at any first-line visit (40 (20%) at one visit only). Each retreatment visit was missed by at most 4 (6%) participants, with the exception of week-8 post-retreatment (missed by 10 (16%) participants). Self-reported non-adherence was substantially higher on retreatment (**Figure S4(c)(d)**, 12-19% across visits, 24 (39%) at any retreatment visit, 17 (27%) at one retreatment visit only).

**Figure 2.**
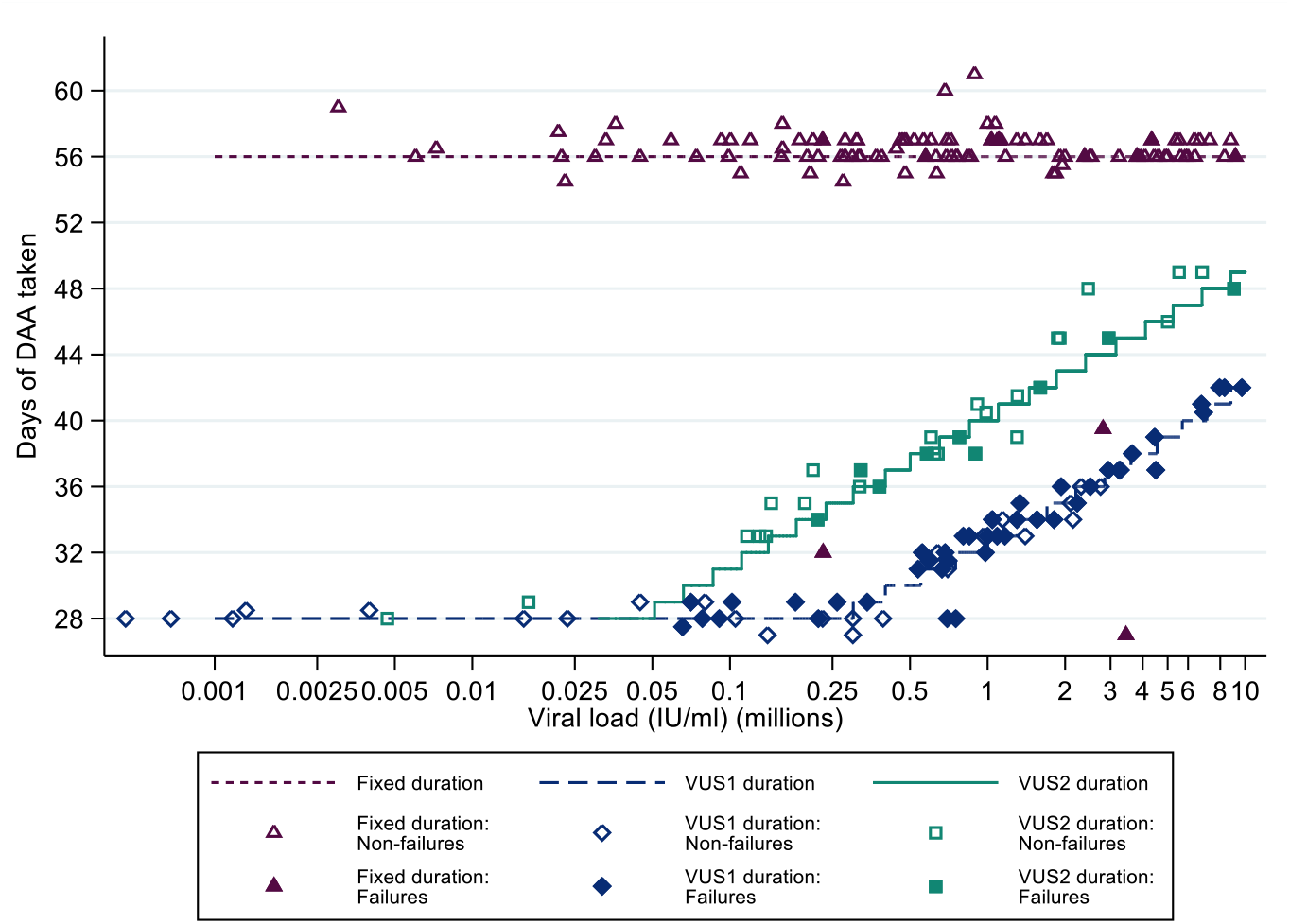
Self-reported treatment duration and first-line SVR24. Note: Lines represent protocol determined treatment duration according to screening viral load (x axis). Symbols represent self-reported individual duration of therapy. Patients could stop DAAs early for adverse events or personal reasons, and take DAAs for longer than prescribed if any missed doses were taken at the end of treatment. Excludes one patient lost-to-follow-up during first-line whose outcome on first-line is unknown.

### Sustained Virological Response (SVR)

Details of the ascertainment of SVR12 are located in **Supplementary Results**. Overall, all evaluable participants achieved SVR12 (and SVR24) on first-line plus retreatment (primary outcome for fixed vs variable-duration randomisation) (100% (95%CI 96%,100%) in both groups, difference 0% (95%CI (Newcombe) - 3.8%,+3.7%), within the pre-specified 4% non-inferiority margin).

First-line SVR12 (secondary outcome) was 91% (95% CI 86%,97%; 92/101) in the fixed-duration group vs 48% (39,57%; 47/98) in the variable-duration group (**Figure 3(a)**; difference −43% (95% CI −54%,-32%), p<0.0001). However, SVR12 was significantly higher for VUS2 (72% (56%,87%); 23/32) than VUS1 (36% (25%,48%); 24/66) (interaction between duration randomisation and strategy p=0.001). First-line SVR12 was 72% (65%,78%); 70/101) with ribavirin and 68% (61%,76%; 69/98) without (difference −3% (−13%,+6%) p=0.48 adjusting for interaction between duration randomisation and strategy). There was no evidence of interaction between ribavirin and duration randomisations overall (heterogeneity p=0.16 adjusted for the interaction between duration randomisation and strategy) or in the variable-duration group, where SVR12 was 52% (37%,67%; 25/48) with ribavirin vs 44% (30%,59%; 22/50) without ribavirin (difference 8% (95% CI −10%,+27%), p=0.38).

**Figure 3.**
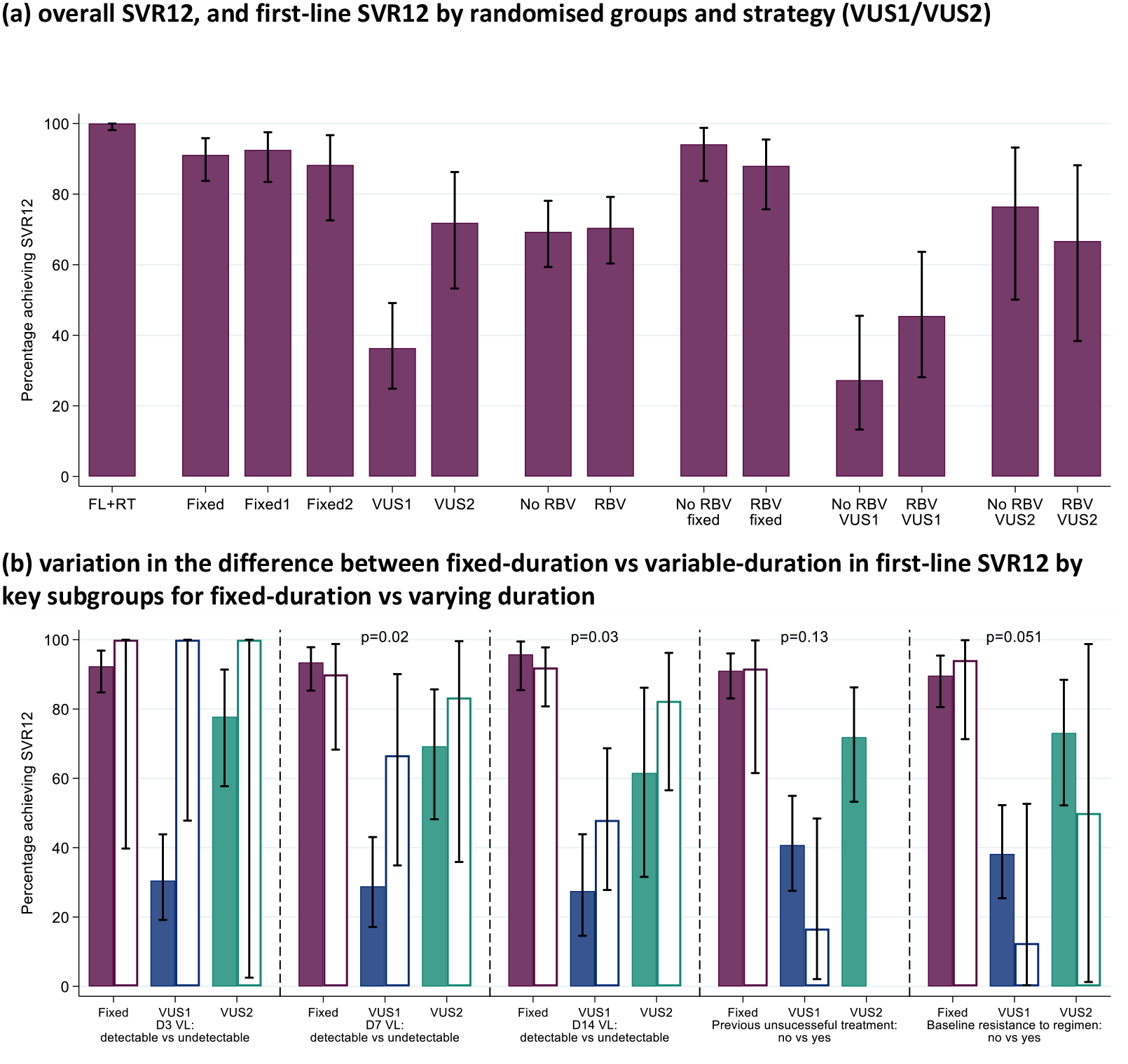
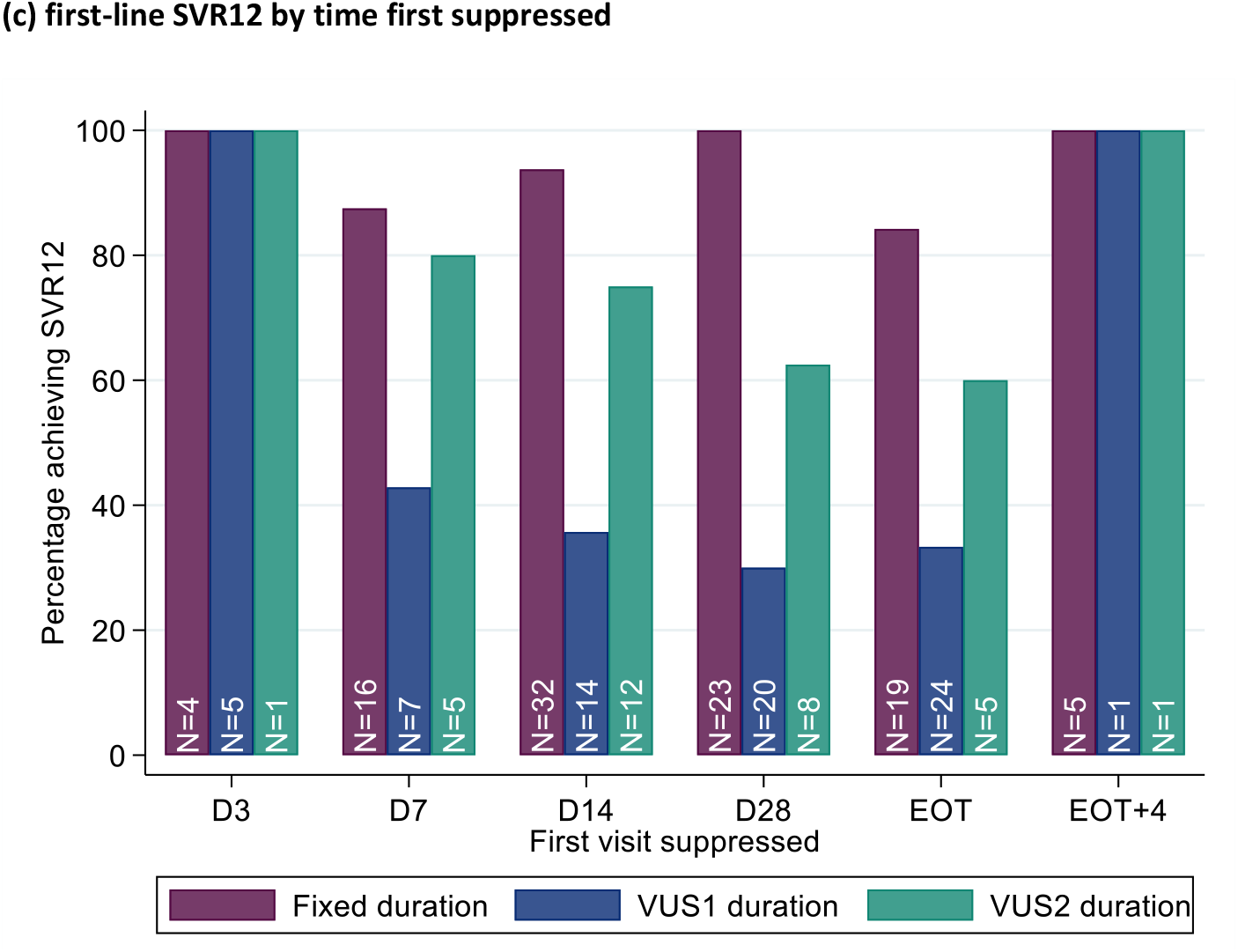
SVR12. Note: FL=first-line, RT=retreatment. Fixed = overall SVR12 for 8 week therapy. Fixed1=fixed duration when VUS duration received VUS1, Fixed2=fixed duration when VUS duration received VUS2. In panel (b), solid bars represent the first subgroup (detectable VL at the various days shown, no previous unsuccessful treatment, no baseline resistance to drugs taken first-line) and empty bars the second subgroup (undetectable VL at the various days shown, previous unsuccessful treatment, baseline resistance to drugs taken first-line). p-values are heterogeneity p-values comparing the difference between fixed vs VUS1/VUS2 strategies across the two subgroups. Heterogeneity p-value for D3 VL cannot be estimated due to perfect prediction. Panel (c) does not include 8 patients who never had VL suppression; for patients allocated 28-31 days, their EOT visit is also their 28 day visit and they are included in each group.

The difference in first-line SVR12 between fixed-duration vs variable-duration was significantly smaller in 2 of 16 subgroups pre-specified in the protocol, suppression at day-7 or day-14 (heterogeneity p=0.02, 0.03 respectively) (**Figure 3(b), Figure S5**). Considering the time when individuals first became undetectable, all 10 individuals who became undetectable at day-3 of treatment achieved first-line SVR12 regardless of treatment duration (as did 31/38 (82%) of those first undetectable at day-7) (**Figure 3(c)**).

Results were similar in the per-protocol population which comprised 142 (70%) participants (most exclusions due to differences between screening and enrolment VL) and in other sensitivity analyses (**Supplementary Results**).

### Emergence of resistance

14 participants who failed on first-line treatment developed a new resistance mutation (not present at baseline) to at least one of their prescribed drugs (**Tables S5, S6**). Within paired samples available at baseline and failure, there was no evidence of a difference in emergent resistance between fixed-duration and variable-duration (3/10 (30%) vs 11/46 (24%, respectively, p=0.77). However, ribavirin was associated with lower emergence of resistance to first-line drugs (3/27 (11%) with vs 11/29 (38%) without ribavirin, p=0.01; **Figure 4**) with significantly lower rates of resistance to any DAA and to NS5a inhibitors (both p=0.01; classified by retreatment drugs in **Figure S7**).

**Figure 4.**
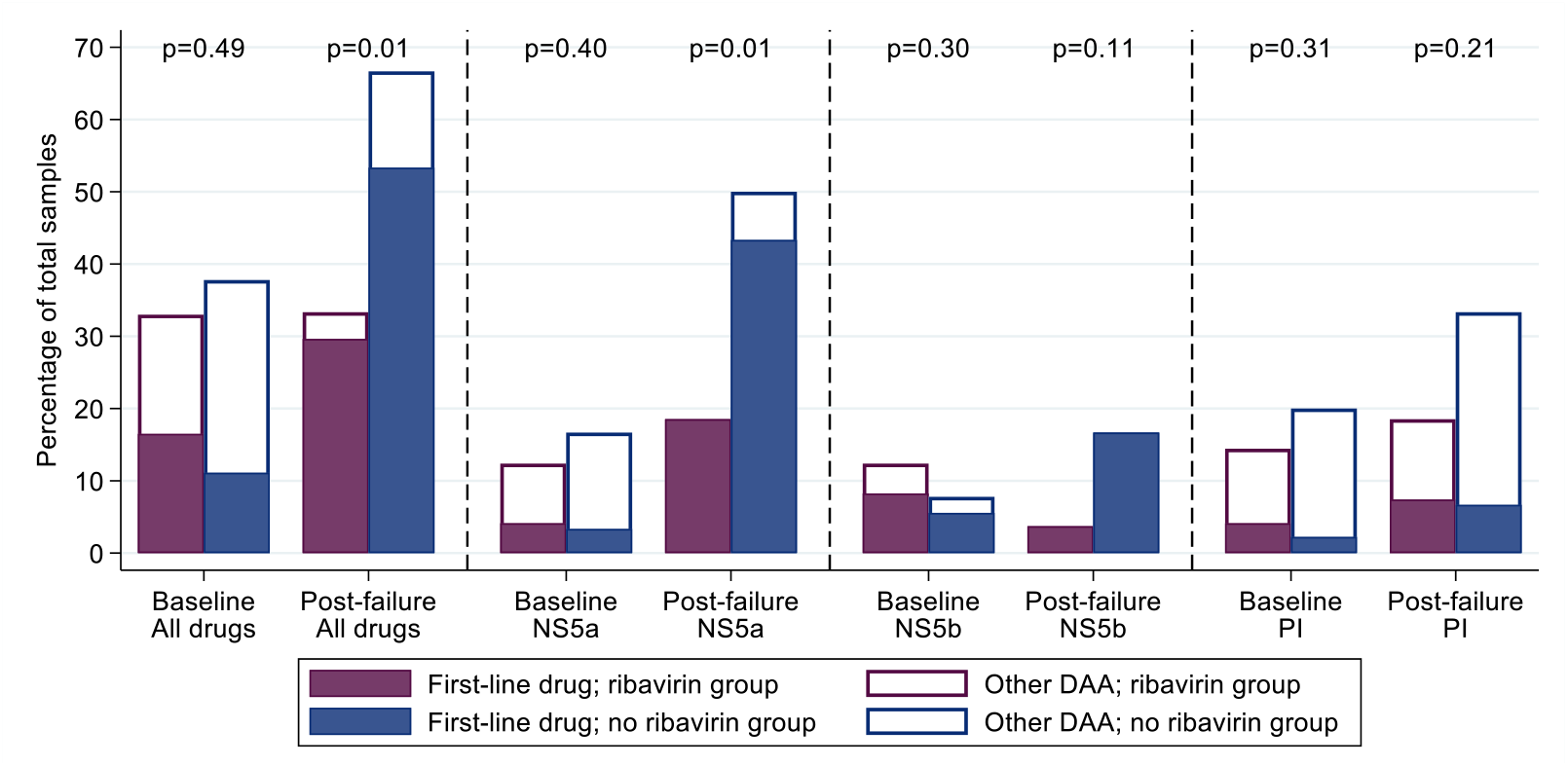
Resistance at baseline (all tested participants) and at first-line failure (failures only) Note: DAA: direct acting antivirals. p-values are comparing any resistance, to first-line drug or any other DAA, between ribavirin groups. See **Figure S7** for resistance classified according to retreatment drugs. Coloured bars represent resistance to drugs received first-line, white bars, all DAAs including those not used in the trial.

### Adverse events

Overall, 5 (5%) variable-duration vs 5 (5%) fixed-duration participants experienced SAEs (hazard ratio (HR)=0.77 (95% CI 0.21,2.80) p=0.69) and 9 (9%) vs 5 (5%) respectively experienced Grade 3/4 AEs (HR=1.74 (0.58,5.24) p=0.33) (**Tables S7, S8**). Similarly, 5 (5%) ribavirin vs 5 (5%) no ribavirin participants experienced SAEs (HR=1.05 (95% CI 0.30,3.63) p=0.59) and 9 (9%) vs 5 (5%) respectively experienced Grade 3/4 AEs (HR=1.92 (0.64,5.72) p=0.59) (**Tables S8, S9**). Treatment-related AEs, AEs causing changes to treatment and Grade 3/4 anaemia were all uncommon (**Table S7**); all Grade 3/4 anaemias occurred in participants randomised to adjunctive ribavirin (p=0.12) as did all first-line drug changes for AEs (p=0.06) (**Table S9**).

## DISCUSSION

This large strategic post licensing trial demonstrated overall non-inferiority of strategies using first-line ultrashort treatment durations, with both groups achieving 100% SVR12 rate after retreatment. The initial shortening strategy (VUS1) was able to cure only 36% of participants first-line but, strikingly, a relatively small increase in ultrashort treatment duration (from a mean of 32 days to 39 days) resulted in a doubling of SVR12 rates (from 36% to 72%). The 8-week fixed-duration strategy, with an SVR12 of 91%, did not have higher efficacy than previous phase II trials of shorter treatment courses,^17^ despite limiting the maximal baseline viral load in those enrolled, which might have been expected to reduce the risk of failure. The trial’s findings suggest a high proportion of patients can be cured with, on average, approximately 60% of the licensed duration of first-line therapy with the agents used in the trial (VUS2 strategy). However, first-line cure rates were not sufficient to be routinely recommended for stable patients able to adhere to 8-12 weeks’ therapy.

Previous work has found adherence to DAA therapy declines as treatment progresses, with patients citing “feeling as if the treatment is working” as a reason for decreasing adherence.^5^ Despite trial participants generally being considered to have better adherence, we found similarly decreasing adherence with time on first-line, with 28% reporting missed first-line doses, and poorer adherence to retreatment (despite its 100% SVR12) (**Figure S4**). That we observed 72% SVR12 with VUS2 (mean duration 39 days) despite 28% reporting missing doses, suggests that intermittent non-adherence may be less important than overall adherence during weeks 4 to 8 of first-line treatment, and emphasise the importance of supporting adherence after week 4 of therapy to ensure good cure rates in hard-to-reach populations.^10,18^

The risk of virological failure, in a patient unlikely to complete the recommended treatment course, is a clinical concern. Emergent resistance could compromise retreatment, particularly where retreatment does not include a protease inhibitor (as in this trial, in contrast to licensed retreatment options). This is also an ethical consideration in short-course therapy trials. We found the first-line treatment strategy did not compromise participants’ ability to ultimately achieve SVR12. Whether this would be the case with pan-genotypic first-line treatment regimens remains to be tested, although it seems plausible. The 100% SV12 rates for retreatment is reassuring from an ethical perspective and suggests that, in certain circumstances, the combination may represent a viable retreatment option for patients failing therapy where access to licensed retreatment options (such as sofosbuvir/velpatasvir/voxilaprevir) remains limited.

Although ribavirin side-effects, particularly anaemia and fatigue, increasingly limit its use ^2,12^ it still has a role for some patients^13^ with limited evidence that it can increase efficacy^19^ and in vitro evidence that it may reduce the emergence of resistance with short course therapy.^20^ Here, additional ribavirin was well tolerated, with only 2-4% participants experiencing adverse events. Across randomised groups, there was no evidence of improvement in SVR12. However, the emergence of resistance was significantly lower in those failing therapy (12% with ribavirin v 38% without), the first time this has been demonstrated in a randomised trial. This suggests that adjunctive ribavirin may have a role for some patients considered at high risk of not completing therapy in order to reduce the risk of compromising retreatment and further work is required to understand better the mechanism of action in this setting.

Response-guided therapy, shortening treatment based on initial virological response, was commonplace for interferon-based therapy ^21^ but this approach is not currently recommended for DAA therapy.^22,23^ Our findings suggest, for the first time, that very early responses to treatment (undetectable at day-3, or even day-7) may be helpful in predicting success of shortened treatment courses. Whilst not widely applicable, in specific supervised clinical settings (including in-patients, prisoners or directly observed daily therapy in the community) such an approach may help guide management and deserves further confirmation in prospective studies^23,24^ .

This trial was designed to test treatment strategies, rather than specific regimens. Almost all recruitment happened when ombitasvir, paritaprevir, dasabuvir and ritonavir (Viekirax, Abbvie) was the preferred first-line treatment in the UK National Health Service (NHS). This combination remains a recommended NHS option, part of the WHO Essential Medicines List and is used in a number of countries. However, its used has been superceded by pangenotypic options in many settings and the extent to which these findings can be generalised to other combinations with broader genotype coverage is unknown.

Give the similar declines in HCV VL between this and other DAA combinations (**Figure S10**), it seems plausible that the relationship between treatment duration and SVR12 is similar for other DAAs combinations approved for 12 weeks’ for patients with mild disease.

The trial required a population able to adhere to a schedule with significantly more visits than standard of care. Non-attendance was low and self-reported adherence reasonably high (**Figure S4**), despite one-third participants actively using recreational drugs. Following expanded access to DAA therapy, trial recruitment completed short of its original target when there were very few patients in need of treatment able to adhere to the follow-up schedule. However, higher than anticipated success of retreatment (predicted to be 85%, actually 100%) and lower than expected loss to follow-up meant that the trial was able to demonstrate non-inferiority according to its pre-specified margin, providing confidence that either strategy would result in an overall SVR12 rate of at least 96% (higher than originally specified).

The treatment of individuals who are unlikely to complete recommended treatment courses is crucial for elimination strategies. Our findings suggest that ultrashort-courses of treatment can cure a significant proportion of patients with mild liver disease, without compromising retreatment in those not cured. Additional ribavirin in those unlikely to complete a course of treatment may be helpful to prevent the emergence of resistant virus.

## Data Availability

Data sets related to this trial are available on application to the STOPHCV trial management committee

## ACKNOWLEDGEMENTS

We thank all the participants and staff from all the centres participating in the STOP-HCV-1 trial (listed in supplementary material) and NHS England specialist commissioning. We thank the Trial Steering Committee (Professor Ian Weller (Chair), Dr Patrick Ingiliz, Dr Colette Smith, and Mr Robert James (Public and Patient representative)) and the Data Monitoring Committee (Professor David Lalloo (Chair), Professor Tim Peto, Professor Katherine Fielding and Dr Oscar Bortolami).

## Authors contributions

GC was the chief investigator. The trial was conceived and designed by GC, ASW, EB, SLP. The trial was managed by GC, ASW, SLP, FH, ED. The site principal investigators, responsible for participant recruitment and data collection, were RG, SV, GC, SR,JC,SB,AA,SB,MN, CC, BS,MW,DF,SM.

Statistical analysis was performed by LMc. Sequencing and bioinformatic analyses were performed by DN, DS, AA. The first draft was written by GC and ASW with review and comment from all authors.

## Figure legends

Figure 1 Trial profile

Note: EOT+12: 12 weeks after the end of treatment; EOT+24 24 weeks after the end of treatment. Some patients lost to follow up are included in the analysis based on local VLs from clinics.

Figure 2 Self-reported treatment duration and first-line SVR24

Note: Lines represent protocol determined treatment duration according to screening viral load (x axis). Symbols represent self-reported individual duration of therapy. Patients could stop DAAs early for adverse events or personal reasons, and take DAAs for longer than prescribed if any missed doses were taken at the end of treatment. Excludes one patient lost-to-follow-up during first-line whose outcome on first-line is unknown.

Figure 3 SVR12

(a) overall SVR12, and first-line SVR12 by randomised groups and strategies (VUS1/VUS2)

(b) variation in the difference between fixed-duration vs variable-duration in first-line SVR12 by key subgroups

(c) first-line SVR12 by time first suppressed

Note: FL=first-line, RT=retreatment. Fixed = overall SVR12 for 8 week therapy. Fixed1=fixed duration when VUS duration received VUS1, Fixed2=fixed duration when VUS duration received VUS2. In panel (b), solid bars represent the first subgroup (detectable VL at the various days shown, no previous unsuccessful treatment, no baseline resistance to drugs taken first-line) and empty bars the second subgroup (undetectable VL at the various days shown, previous unsuccessful treatment, baseline resistance to drugs taken first-line). p-values are heterogeneity p-values comparing the difference between fixed vs VUS1/VUS2 strategies across the two subgroups. Heterogeneity p-value for D3 VL cannot be estimated due to perfect prediction. Panel (c) does not include 8 patients who never had VL suppression; for patients allocated 28-31 days, their EOT visit is also their 28 day visit and they are included in each group.

Figure 4 Resistance at baseline (all tested participants) and at first-line failure (failures only)

Note: DAA: direct acting antivirals. p-values are comparing any resistance, to first-line drug or any other DAA, between ribavirin groups. See **Figure S7** for resistance classified according to retreatment drugs. Coloured bars represent resistance to drugs received first-line, white bars, all DAAs including those not used in the trial.

