## Supplemental files for "Strategic Treatment Optimization for HCV (STOPHCV1): a randomised controlled trial of ultrashort duration therapy for chronic hepatitis C"

**Supplementary material**

### List of investigators

**The STOP-HCV-1 trial group consists of:**

**Participating Centres**:

**Brighton, Royal Sussex County Hospital**: Sumita Verma, Allison Leslie, Maggie Cole, Wezwei Li, Jane Lyttle, Heidi Barkhordar, Mary Flowerdew, Michelle Villanueva, Samantha Aplin, Lisa Furnival, Raquel Nsuesha Akieme, Lorraine Shah-Goodwin, Vicky Kennard, Lucia Macken, Caroline McPherson, Memory Kamhuka, Mel Smith, Michelle Powell, John Bell, Onyinyechi Uwasomba, Dominika Wlazly, Abraham Gihawi, Rhea Savvidov Strudwick, Paul Frattaroli, Ahmed Hashim, Zhengmai He, Christine Laycock, Sephora Shaw, Hannah Heron, Tenesa Sargent

**Edinburgh, Western General Hospital**: Clifford Leen, Sheila Morris, Amy Shepherd, Aileen Cairns, Hazel Rae, Ruaridh Buchan

**Glasgow Royal Infirmary**: Stephen Barclay, Dominic Rimmer, Janet Johnstone, John Alexander, Joan Blevings, Neil Lachlan, Ewan Forrest, Sharon Grant, Susanne Cathcart, Dario Salutous

**Guildford, Royal Surrey County Hospital**: Aftab Ala, Shahrzad Shahmehri, Rebecca Wills, Fiona Butler, Laura Gordon, Jenneke Schmit, Tineke Edmunds, Catherine Medcalf, Veronica Davies, Louise Gallagher, Clare Kelly, Cassila Peixoto, Sanja Mathew , Federica Regis, Lee Moore, Victoria Westall , Max Wong, Nichola Wakeford, Sally Bradfeld, Marinos Pericleous

**Leicester Royal Infirmary**: Martin Wiselka, Adam Lewszuk, Valerie Renals, Kate Ellis, Karl Hastings, Paul Underwood, Natasha Patel

**London, Chelsea & Westminster Hospital**: Mark Nelson, Carina Bautista, Mini Thankachen, Kathryn McCormick, Helen Morgan, Francesca Ferretti, Daniel Bradshaw, Nicole Pagani, Maria Elena Noval, Maddalena Cerrone

**London, Mortimer Market Centre**: Richard Gilson, Sarah Pett, Nicola Stewart, Holly Wing, Marzia Fiorino, Diarmuid Nugent, Binta Sultan, Kristian Warnes, Erica Pool, Ana Milinkovic, Asma Ashraf, Jack Brophy, Lewis Haddow, Hinal Lukha, June Minton, Indrajit Ghosh , Nina Bason

**London, Royal Free Hospital**: Sanjay Bhagani, Anne Carroll, Glykeria Pakou, Filippo Ferro, Jonathan Edwards, Nnenna Ngwu, Alice Nightingale, Thomas Fernandez, Eric Witele, Vivian Wu, Janet North, Mathew Wright, Fatema Essaji, Parizade Raymode, Tabitha Mahungu , Stephanie Harris, Nomusa Mpofu, Conor Bowman, Alison Rodger, Sabina Melander, Aarti Nandani

**London, St. George's Hospital**: Daniel Forton, Hannah Rine, Nick Tatman, Beverly Edwards, Alice Dainty, Helen Webb, Nia Al-Samarrai, Anet Soubieres

**London, St. Mary's Hospital**: Graham Cooke, Christopher Jones, Wilbert Ayap, Chi-man Wong, Ajerico Del Rosario, Michael Wood, Ken Legg, Claire Petersen, Heather Lewis, Andrew Lovell, Maryam Khan

**Newcastle, Freeman Hospital**: Stuart McPherson, Lorna Brownlee, Sarah Hogg, Maureen Foreman, Afnaan Nadeem , Steven Masson, Jack Oliver, Jennifer Gallagher, Shion Gosrani, Helen Dixon, Wendy Nichol, Penny Bradley, Rachael Forbes, Hannah Stevenson, Heather Russell, Phillipa Thwaites, Jade Davison

**Nottingham, Queens Medical Centre**: Stephen Ryder, Juliette Verheyden, Zoe Rose, Jack Squires, Sheila Hodgson, Bernie Cook, Sally Hodgkinson, Paul Douglas, Victoria Scott-South, Elizabeth Keddie Gray, David Nash, Robert Scott , G. Aithal , J. Chalmers, W. Fateen , L. James , M. Barnes , Varinder Ryan, B. Reynolds , R. Harris , P. Thiagarayan , Martin James, Susan Congreave

**Oxford, John Radcliffe Hospital**: Jane Collier , Denise O'Donnell, Lizzie Stafford, Louise Holland, Ellie Barnes, Nellia Sande, Christine Tsang, Wendy Chau, Mark Ainsworth, Anna Simpson, Katerina Klimova, Paola Cicconi, Adele Melton, Oliver Duncan, Evelyn Chan, Forzia Mushtaq, Toby Cade

**Sheffield, Royal Hallamshire Hospital**: Ben Stone, Dr Christopher Durojaiye, Sarah Berry, Deborah Allen, Meena Chopra, Carol Jaques, Tracey Jackson, Helen Bowler, Lynsey Jones, Lynne Smart, Sophia Fell, Esme Marshall, Dr Katherine Cartwright, Dr Alicia Vedio, Susan Thompson, Dr Paul Collini, Cecilia Tawiah

**Swansea, Singleton Hospital**: Chin Lye Ch'ng, Caradog Thomas, Sue Thomson, Lynda Connor , Glyn Gainard, Euan Pratt, Keith Baldwin, Paul John, Helen Thompson-Jones, Sally Kneath

**Trial Coordination and Oversight - MRC CTU at UCL, London**: Ann Sarah Walker, Fleur Hudson, Emily Dennis, Cara Purvis, Leanne McCabe, Sarah Pett, Helen Ainscough, Ania Spurdens, Deborah Alawode, Aminata Sy, Nafisah Atako, Charlotte Russell, Will Everett. **Data Management Systems**: Mary Rauchenberger, Chiara Borg.

**Trial Steering Committee:** (Independent) Ian Weller (Chair), Patrick Ingiliz, Colette Smith, Robert James (Team) Graham Cooke, Ann Sarah Walker, Eleanor Barnes.

**Data Monitoring Committee**: David Lalloo (Chair), Tim Peto, Katherine Fielding, Oscar Bortolami.

### Supplementary Methods

#### (a) Full inclusion and exclusion criteria

Inclusion criteria

1. Aged ≥18 years
2. Infected with HCV genotype 1a or 1b or 4 with access to first-line treatment appropriate for their genotype (ombitasvir/paritaprevir/(dasabuvir)/ritonavir or glecaprevir/pibrentasvir)
3. At least one detectable viremia 6 months prior to randomisation (by quantitative HCV RNA, qualitative assay or HCV genotype), with no intervening undetectable results
4. Plasma HCV RNA >lower limit of quantification at screening
5. No evidence of significant liver fibrosis resulting from any aetiology (defined as Fibroscan score ≤7.1kPa, equivalent to F0-F1, within 180 days prior to planned randomisation or biopsy consistent with mild fibrosis (Ishak score <=2/6) within 180 days prior to planned randomisation)
6. BMI >=18kg/m^2^
7. Laboratory tests: platelets >=60x10^9^/L, haemoglobin >12g/dL (male) or >11g/dL (female), creatinine clearance (estimated using Cockcroft-Gault) >=60ml/min, international normalised ratio (INR) <1.5
8. Screening HCV viral load <10,000,000IU/ml
9. Written informed consent obtained from the patient.
10. If HIV infected, then on antiretroviral therapy with HIV viral load <50 copies/ml for >24 weeks at the screening visit.

Exclusion criteria

1. Previous DAA exposure for this infection (previous treatment with pegylated-interferon and/or ribavirin allowed. DAA treatment for a previously cured infection allowed).
2. FEMALES ONLY: Lactating, or pregnant, or planning to become pregnant, or not willing to use effective contraception, during the study and for four months after last dose of study medication.
3. FEMALES ONLY: currently taking ethinyl-oestradiol-containing medicinal products such as those contained in most combined oral contraceptives or contraceptive vaginal rings.
4. MALES only: planning pregnancy with female partner, or not willing to use effective contraception, during the study and for seven months after last dose of study medication.
5. Malignancy within 5 years prior to screening
6. Any condition in the judgement of the investigator which might limit the patient’s life expectancy
7. Currently receiving medication know to interact with study medication (ombitasvir, paritaprevir, dasabuvir, ritonavir, sofosbuvir, ledipasvir, ribavirin, glecaprevir, pibrentasvir; based on relevant prescribing information and www.hep-druginteractions.org)
8. Disorder which may cause ongoing liver disease including, but not limited to, active hepatitis B, ongoing alcohol misuse
9. Any disorder which in the opinion of the investigator may have a significant negative impact on the ability of the patient to adhere to the trial regimen
10. Use of other investigational products within 60 days of screening
11. Known hypersensitivity to any active ingredient and/or excipients of the study medicines, namely Microcrystalline cellulose, Lactose monohydrate, Croscarmellose sodium, Magnesium stearate, Gelatine, Shellac, Propylene glycol, Polyethylene glycol, Ammonium hydroxide, Pregelatinised maize starch, Sodium starch glycolate (type A), Maize starch, Hypromellose, Talc, Ethylcellulose aqueous dispersion, Triacetin, Copovidone, Colloidal anhydrous silica, vitamin E (tocopherol) polyethlyene glycol succinate, sodium stearyl fumarate, Polyvinyl alcohol, Macrogol 3350, Sunset yellow FCF aluminium lake (E110), Colouring agent (E132), Titanium dioxide (E171), Yellow iron oxide (E172), Red iron oxide (E172), Black iron oxide (E172).
12. History of severe pre-existing cardiac disease, including unstable or uncontrolled cardiac disease, in the previous six months
13. Haemoglobinopathies (e.g., thalassemia, sickle-cell anaemia).

Further details of randomisation

Randomisation used a minimisation algorithm incorporating a probabilistic element incorporated securely into the online trial database. Allocation was concealed until eligibility was confirmed by researchers at local hospitals; randomisation was then performed centrally.

#### (b) Further details of endpoint ascertainment

Serious adverse events (SAEs) were defined following the International Committee for Harmonization as events which led to death, were life-threatening, caused or prolonged hospitalisation (excluding elective procedures), caused permanent disability, or were other medical conditions or with a real, not hypothetical risk of one of the previous categories.

#### (c) Statistical methods

Analyses followed the principle of intention-to-treat including all follow-up regardless of changes to treatment. The Statistical Analysis Plan (SAP) pre-specified that any patient who was randomised in error (defined as realising that the patient should not have been randomised before taking study drug and not ever taking study drug) and hence not followed up would be excluded.

Lost to follow-up (LTFU) was formally defined as not having been seen at the final EOT+24 week visit within a [-6, +12] week window. If a patient was LTFU before the visit at which an outcome was measured (EOT+12 for SVR12, EOT+24 for SVR24), the following methods were pre-specified in the SAP, but not the protocol, to be used to determine the patient’s outcome:

- If the missing HCV VL was between two undetectable measurements, it was assumed to be undetectable
- HCV VL results from local practice were sought for patients with missing SVR12 and SVR24 outcomes. If the local result was undetectable, the patient was assumed to be undetectable at EOT+12/EOT+24. (One patient had a detectable local VL, but as they had no confirmatory subsequent VL they were not considered to have failed and were also not counted as a cure.)

After following these methods, the proportion without an outcome was <10% and so the analysis was restricted to complete cases (as pre-specified in the SAP).

Primary analyses of outcomes restricted to first-line therapy were stratified by first-line DAA strategy in place (VUS1 (before 1 April 2017) or VUS2 (after 1 April 2017)) as a main effect, and as an interaction with randomised group (fixed-duration vs variable-duration), where the p-value for the interaction term was <0.05. Primary analyses of outcomes including retreatment were unstratified, reflecting the overall strategy comparison and because no patients failed after receiving retreatment. Primary analyses were not stratified by centre given the large number of centres with small numbers.

Primary analysis of the primary endpoint included all randomised participants other than those considered randomised in error (following the statistical analysis plan) and for whom no viral load data could be obtained. A per-protocol analysis included patients receiving >90% and <100% of the prescribed duration of first-line treatment and where the difference between screening and enrolment HCV RNA values would have led to a difference of ≤2 days in allocated duration of DAAs had they been allocated to the variable-duration group. Secondary analyses were conducted considering all LTFU patients as failures and all LTFU patients as cured. An additional secondary analysis excluding reinfections identified by genome sequencing pre-specified in the protocol was not performed as no reinfections were identified.

For the primary analysis, a risk difference and 95% confidence interval obtained from a binomial regression on the risk difference scale using a generalised linear model. Kaplan-Meier plots and Cox proportional hazard models were used for analyses of time until failure (any type). Secondary analyses of primary treatment failure and viral load rebound (the components of overall treatment failure) used competing risks methods (cumulative incidence plots, subhazard ratios) to account for the possibility of the patient experiencing the other type of failure. Binomial generalised estimating equations with an independent working correlation were used to analyse the proportion of patients with undetectable HCV VL at each time point.

Safety outcomes were analysed using chi-squared p-values. To assess the change in laboratory values over time (other than for HCV VL), generalised estimating equations (normal distribution) with an independent correlation structure adjusted for baseline values were used. Sensitivity analyses of changes in laboratory values used alternative error structures and mixed effects models, but these provided similar results to the primary analysis.

Baseline values of laboratory test results were those taken closest to, but before randomisation. HCV VL was log_10_ transformed for analysis as a continuous variable. Other continuous measures were transformed using Box-Cox transformations when there was gross (p<0.0001) deviations from normality as assessed using the Shapiro-Wilk test. Analyses of measurements at a given point in follow up used the closest available measurement to that time point in evenly spaced windows. If a visit fell in two visit windows, it was classed as belonging to the latter window, except where this led to no visits within in the first window and two within the second, in which case it was classed as belonging to the first visit window.

#### (d) Subgroup analyses

Subgroup analyses were conducted to assess consistency of effects across different participant characteristics. All subgroup analyses were adjusted for the interaction between VUS strategy and duration randomisation due to it being highly significant in the primary analysis. For the duration comparison, interaction tests within binomial models on the risk difference scale were used for subgroup analyses of the duration comparison. For the ribavirin comparison, due to non-convergence of the models, p-values were obtained from marginal effects after logistic regression for subgroups. Heterogeneity p-values for IL28B polymorphisms considering C as an ordinal factor were obtained from ordered logistic regression. Continuous factors were categorised into terciles as well as using fractional polynomial models. Heterogeneity p-values could not be estimated for all subgroups due to small numbers or perfect prediction. No formal adjustment for multiple testing was made for subgroup analyses.

The following pre-specified subgroup analyses were performed:

- duration strategy (VUS1 vs VUS2);
- each of the other randomisations to investigate interactions in the factorial design;
- baseline HCV VL as a continuous interaction;
- baseline HCV VL as a dichotomisation at 6,000,000 IU/ml;
- genotype 1a vs 1b vs 4;
- HIV co-infection;
- IL28 polymorphisms (CC vs CT vs TT) as with C as a nominal factor and C as an ordinal factor;
- IL28 polymorphisms (CC vs CT/TT);
- age;
- sex;
- BMI;
- early HCV RNA treatment responses, to days 3, 7 and 14 (undetectable vs detectable HCV VL);
- previous (failed) treatment with interferon-alpha with/out ribavirin, overall;
- presence of viral quasispecies including resistance.

### Supplementary Results

#### Ascertainment of SVR12

One participant withdrew consent (retreatment week-4) and a further 13 (6%) participants were lost-to-follow-up (1 on first-line, 9 post first-line EOT, 2 on retreatment, 1 post retreatment EOT; total withdrawn/lost 9 (9%) fixed-duration, 4 (4%) variable duration). However, HCV VL results were available from medical notes for most of those not withdrawing consent, meaning first-line SVR12 and SVR24 could not be ascertained for only 3 (1%) and 6 (3%) participants, respectively (1/2 and 3/3 fixed/variable-duration, respectively), and overall (first-line plus retreatment) SVR12 and SVR24 for only 5 (2%) and 8 (4%) participants (2/3 and 4/4 fixed/variable-duration) respectively (see **Supplementary Methods**).

#### (a) Per-protocol analysis of first-line SVR12, duration and ribavirin randomisations

70 (70%) receiving VUS1/VUS2 vs 72 (71%) receiving fixed-duration were included in the per-protocol population and 68 (68%) receiving ribavirin vs 74 (73%) not receiving ribavirin (received >90% and <100% of the prescribed duration of first-line treatment and had a difference between screening and enrolment HCV RNA values leading to a difference of ≤2 days in allocated duration of DAAs had they been allocated to the variable-duration group).

Duration randomisation:

SVR12 after first-line and any retreatment was 100% overall (95% CI 97%, 100%) with a difference of 0% (95% CI (Newcombe) -5%, +5%).

SVR12 after first-line only was 47% (95% CI 36%, 59%; 32/69) in the variable-duration group vs 93% (95% CI 87%, 99%; 66/71) in the fixed-duration group. The difference was -46% (95% CI -59%, -33%; p<0.0001).

Ribavirin randomisation:

SVR12 after first-line and any retreatment was 100% overall (95% CI 97%, 100%) with a difference of 0% (95% CI (Newcombe) -0.06%, +0.05%).

SVR12 after first-line only was 70% (95% CI 61%, 78%; 48/66) in the ribavirin group vs 70% (95% CI 62%, 78%; 50/74) in the no ribavirin group. The difference was -0% (95% CI -11%, +10%; p=0.93).

#### (b) Other sensitivity analyses of first-line SVR12, duration and ribavirin randomisations

All missing SVR12 considered failures.

Duration randomisation:

SVR12 after first-line and any retreatment was 97% (9%% CI 94%, 100%; 97/100) in the variable-duration group vs 98% (95% CI 95%, 100%; 100/102) in the fixed-duration group. The difference was -1% (95% CI -5%, 3%; p=0.64).

SVR after first-line only was 47% (95% CI 38%, 56%; 47/100) in the variable-duration group vs 90% (95% CI 84%, 96%; 92/102) in the fixed-duration group. The difference was -45% (95% CI -54%, -32%; p<0.0001).

Ribavirin randomisation:

SVR12 after first-line and any retreatment was 97% (95% CI 94%, 100%; 97/100) for the ribavirin group vs 98% (95% CI 95%, 100%; 100/102) in the no ribavirin group. The difference was -1% (95% CI -5%, +3%; p=0.64).

SVR12 after first-line only was 67% (95% CI 60%, 75%; 69/100) in the ribavirin group and 71% (95% CI 65%, 78%; 70/102) in the no ribavirin group. The difference was -4% (95% CI -14%, +5%; p=0.37).

All missing SVR12 considered cured.

Duration randomisation:

SVR12 after first-line and any retreatment was 100% overall (95% CI 98%, 100%) with a difference of 0% (95% CI (Newcombe) -3.7%, +3.7% within the pre-specified 4% non-inferiority margin).

SVR12 after first-line only was 49% (95% CI 40%, 59%; 49/100) in the variable-duration group vs 91% (95% CI 86%, 97%; 93/102) in the fixed-duration group. The difference was -42% (95% CI -53%, -31%; p<0.001).

Ribavirin randomisation:

SVR12 after first-line and any retreatment was 100% overall (95% CI 98%, 100%) with a difference of 0% (95% CI (Newcombe) -3.7%, +3.6% within the pre-specified 4% non-inferiority margin).

SVR12 after first-line only was 69% (95% CI 62%, 76%; 71/100) in the ribavirin group vs 72% (95% CI 65%, 79%; 71/102) in the no ribavirin group. The difference was -3% (95% CI -12%, +6%; p=0.48).

#### (c) SVR24, duration and ribavirin randomisations

Duration randomisation:
SVR24 after first-line and any retreatment was 100% overall (95% CI 98%, 100%; 194/194) with a difference of 0% (95% CI (Newcombe) -3.8%, +3.8%).
SVR24 after first-line only was 47% (95% CI 38%, 56%; 46/97) in the variable-duration group vs 89% (95% CI 83%, 95%; 88/99) in the fixed-duration group. The difference was -42% (95% CI -53%, -31%; p<0.0001).

Ribavirin randomisation:
SVR24 after first-line and any retreatment was 100% overall (95% CI 95%, 100%); 194/194) with a difference of 0% (95% CI (Newcombe) -3.8%, 3.8%).
SVR24 after first-line only was 69% (95% CI 61%, 76%; 68/97) in the ribavirin group vs 68% (95% CI 60%, 76%; 66/99) in the no ribavirin group. The difference was 1% (95% CI -9%, +11%; p=0.87).

#### Timing of failure

Only 1 (0.5%) (VUS1) participant failed on treatment (at EOT, 28 days DAAs) (**Figure S8a**). 21 (10%) participants had primary first-line failure (VL never confirmed undetectable) (5 (5%) fixed-duration vs 16 (16%) variable-duration, p=0.008, **Figure S8(c,ii)**); in the variable-duration group, primary failure occurred in 16 (24%) VUS1 vs 0 (0%) VUS2 (p=0.002; interaction not estimable across strategies). 41 (20%) participants had VL rebound after confirmed undetectable HCV VL (6 (6%) fixed-duration vs 35 (35%) variable-duration, p<0.0001, **Figure S8(c,iii)**; 26 (38%) VUS1, 9 (28%) VUS2, heterogeneity p=0.60). There was no evidence that ribavirin was associated with primary failure (p=0.83) or rebound (p=0.59) (**Figure S9**). Failure tended to occur earlier with VUS1 vs VUS2 (p=0.08**, Figure S8a**), and with variable-duration vs fixed-duration (p=0.07; VUS1 vs fixed-duration p=0.03). However, there was no evidence of differences in failure VLs (median 158073 (VUS1), 89125 (VUS2), 346737 (fixed-duration), p(VUS2 vs VUS1)=0.41, p(VUS2 vs fixed-duration)=0.21) (**Figure S8(b)**). All participants who met failure criteria on first-line started retreatment, a median (IQR) 2.9 (2.0,4.4) weeks after failure was confirmed.

Corresponding to the timing of failures, the percentages with undetectable VL decreased more rapidly post-EOT in VUS1 vs VUS2 (p<0.0001) or VUS1 vs fixed-duration (p<0.0001) (**Figure S10**). There was no evidence of differences in the proportions with detectable VL at EOT in VUS1 compared to VUS2 (p=0.33) or fixed-duration (p=1.00). There was no evidence that ribavirin was associated with higher percentages detectable post-EOT overall (p=0.48) or, within the variable-group, between VUS1 vs VUS2 (interaction p=0.17, **Figure S11**).

### Figure S1 Trial schematic

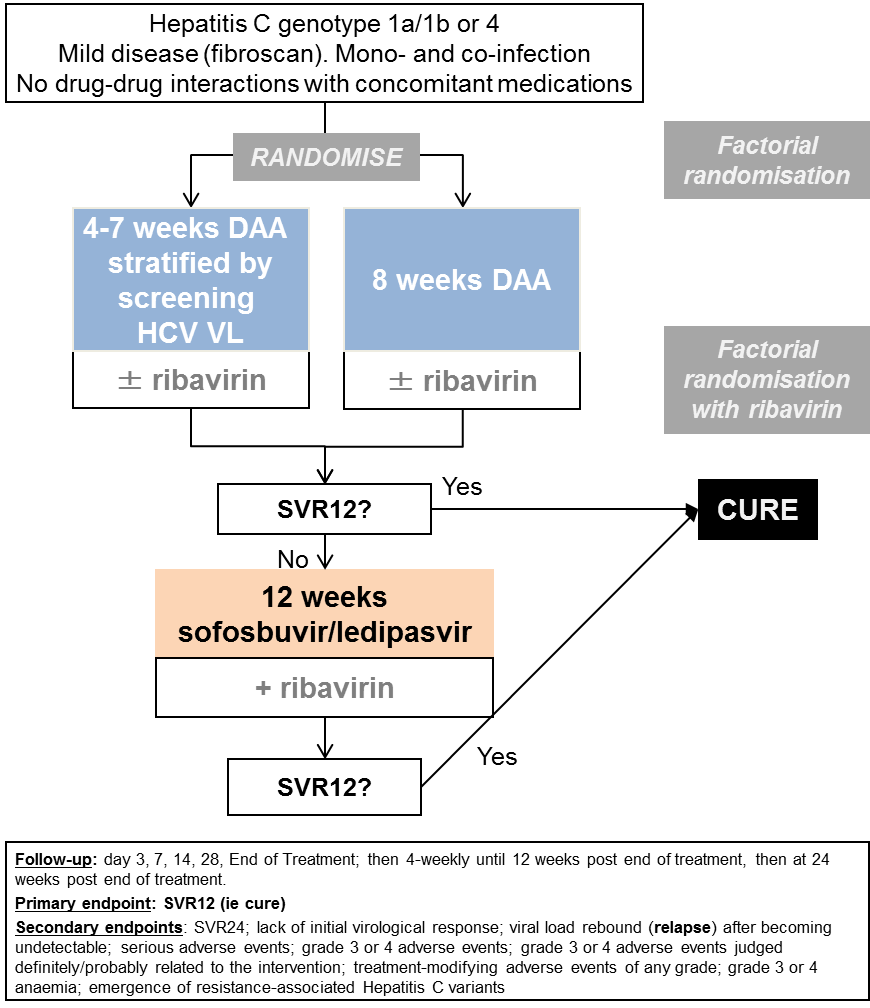

### Figure S2 Duration of first-line treatment in the variable-duration group by protocol version

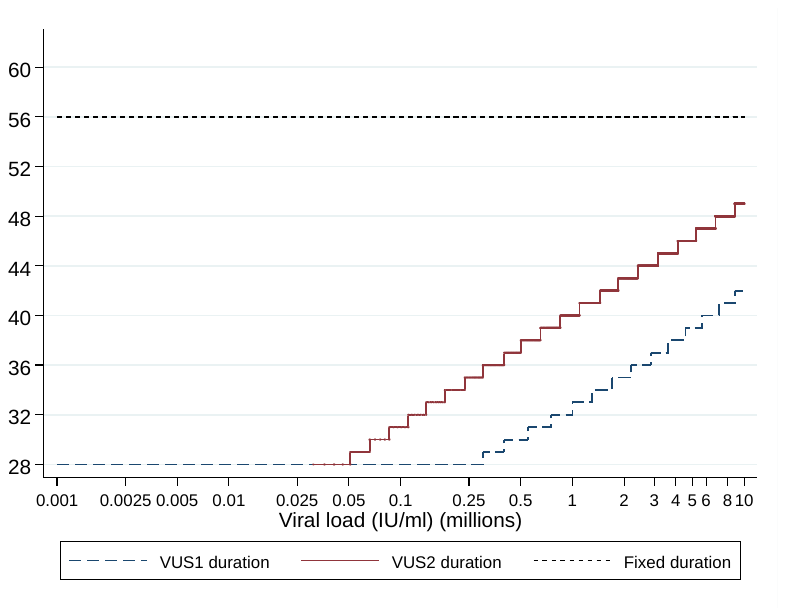

### Figure S3 HCV VL at screening and enrolment (all patients)

#### (a) By assay

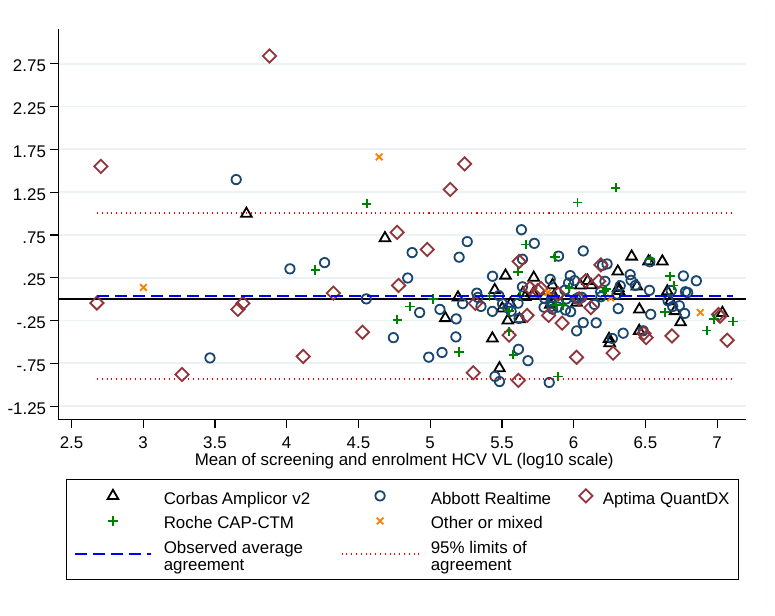

#### (b) By duration randomisation and failure status

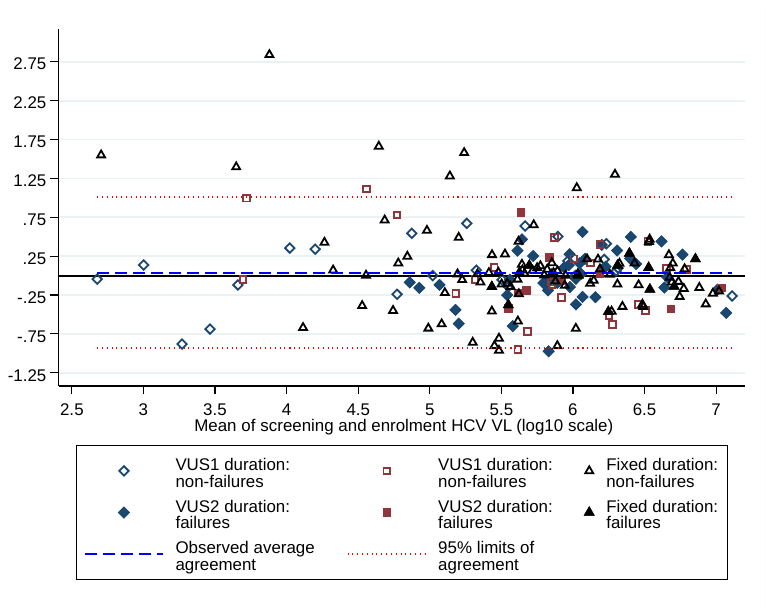

### Figure S4 Reported missing doses (any intervention drug)

#### (a) First-line, by duration randomisation

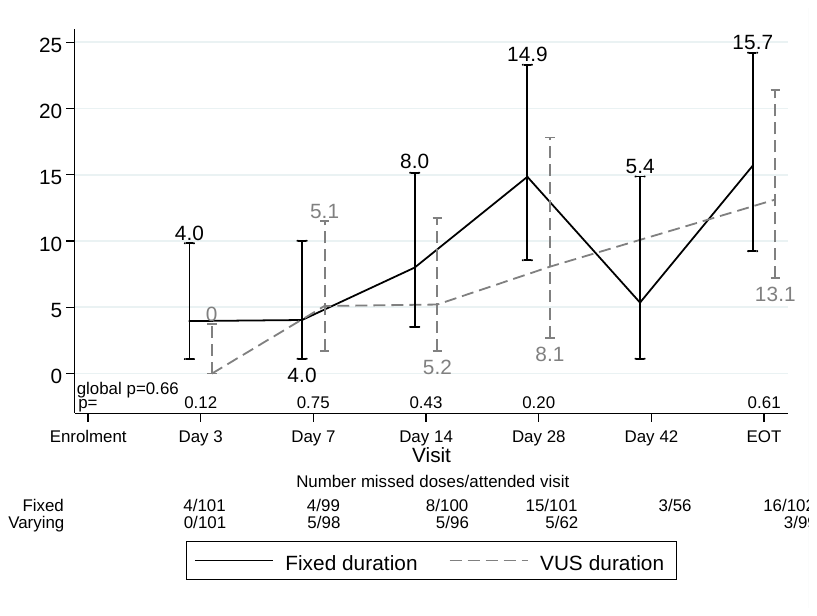

#### (b) First-line, by ribavirin randomisation

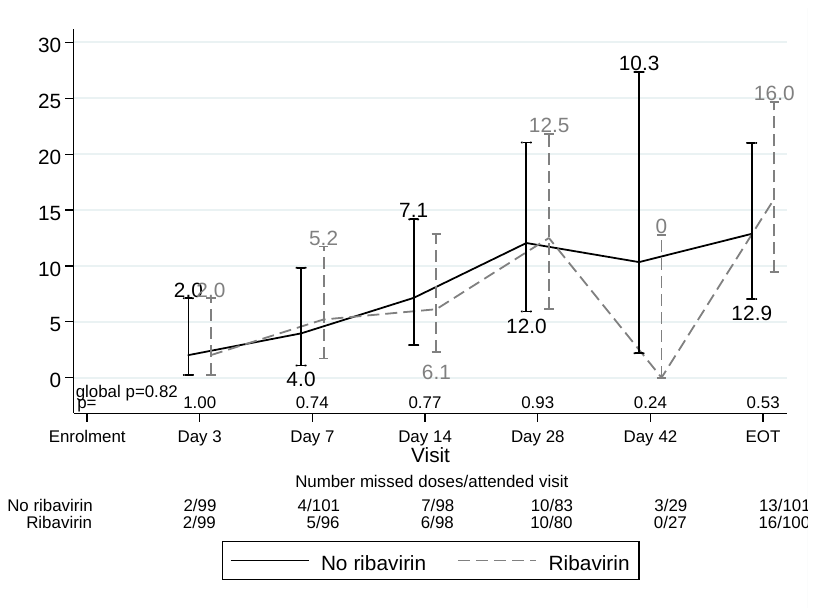

#### (c) Retreatment, by duration randomisation

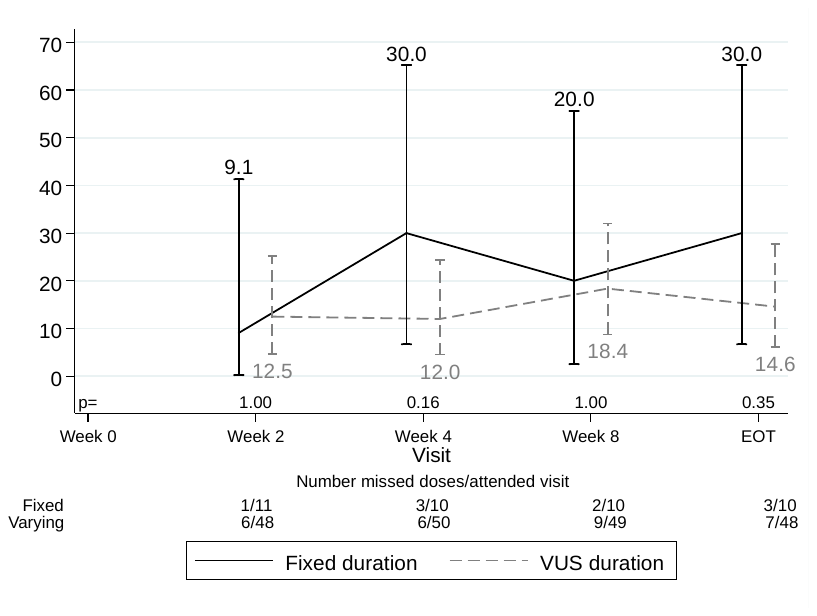

#### (d) Retreatment, by ribavirin randomisation

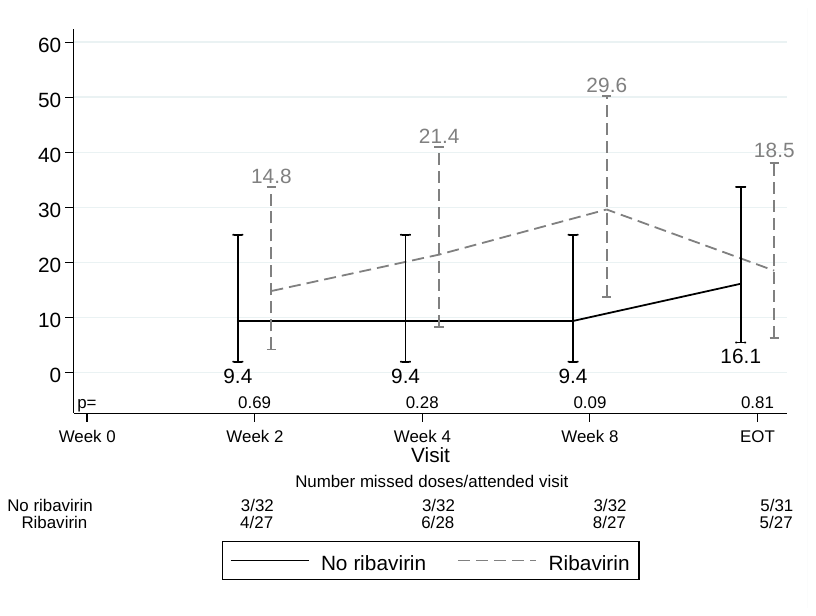

### Figure S5 Subgroup analyses for first-line SVR12 by fixed-duration vs variable-duration randomisation

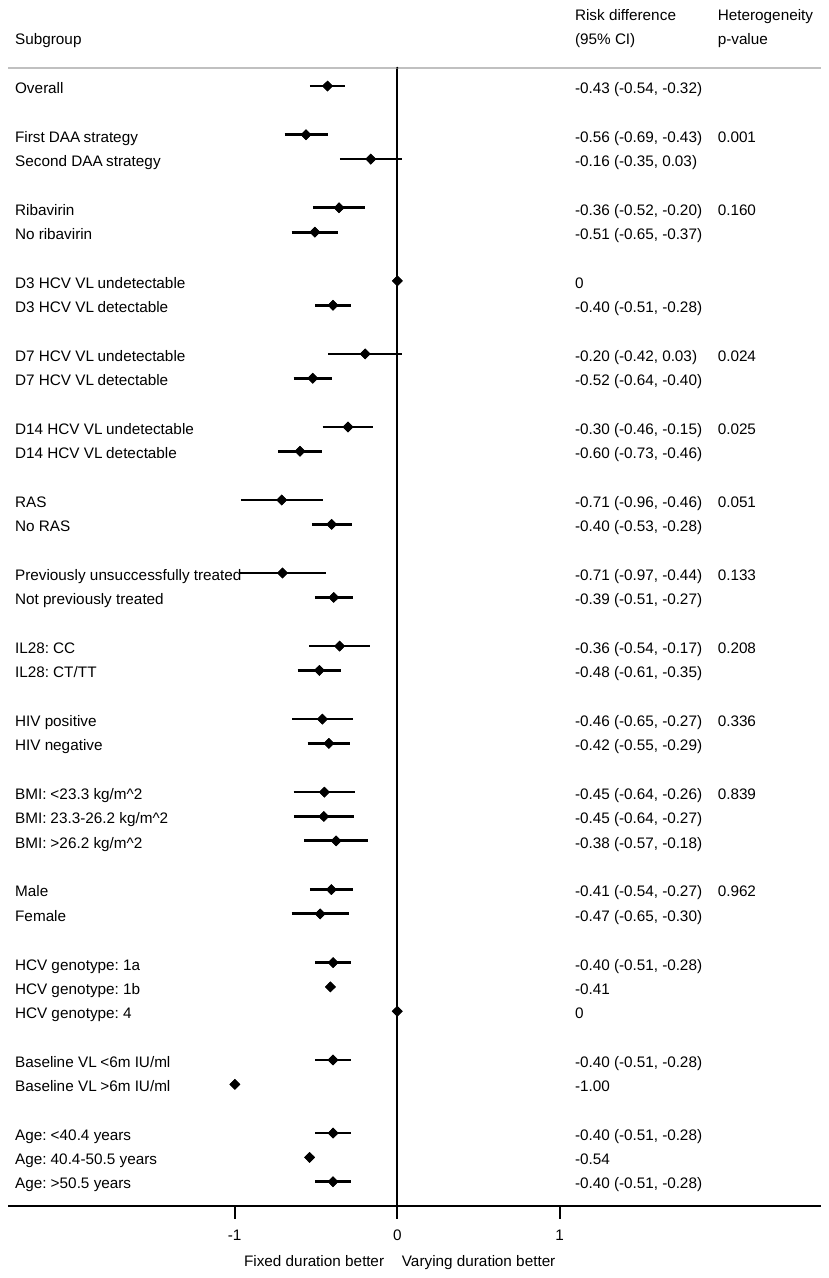

Note: all heterogeneity tests also adjusted for the interaction between strategy and fixed-duration vs variable-duration randomisation. Some confidence intervals and heterogeneity p-values could not be estimated due to perfect prediction.

### Figure S6 Subgroup analyses for first-line SVR12 by ribavirin randomisation

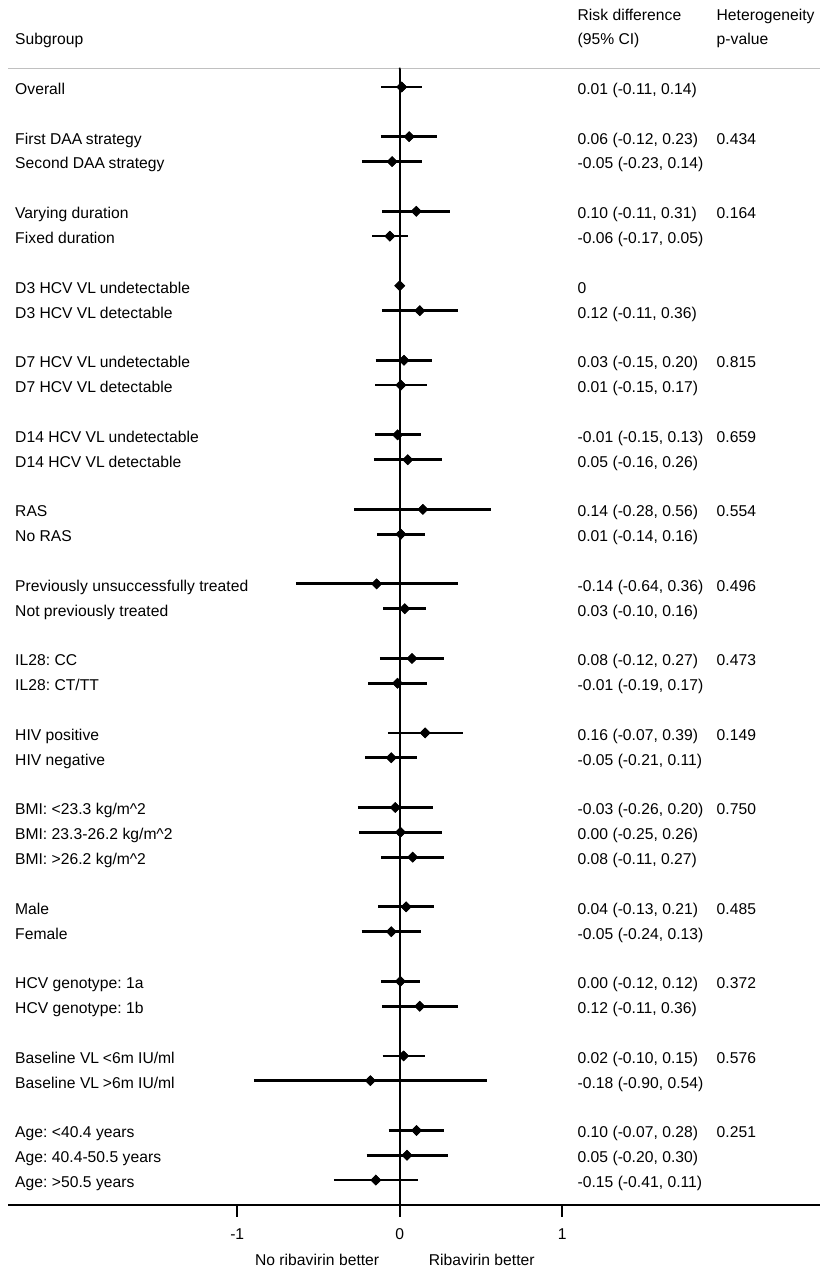

Note: all heterogeneity tests also adjusted for the interaction between DAA strategy and fixed-duration vs variable-duration randomisation. Some confidence intervals heterogeneity p-values could not be estimated due to perfect prediction. All patients with genotype 4 did not receive ribavirin so not included.

### Figure S7 Resistance at first-line failure classified by retreatment or other DAAs

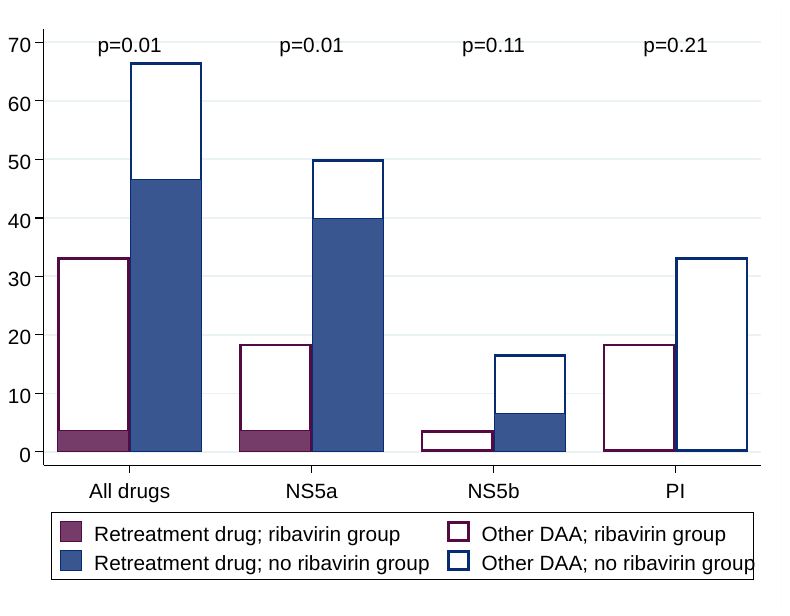

Note: DAA: direct acting antivirals. p-values are comparing any resistance, to retreatment drug or any other DAA, between ribavirin groups. Coloured bars represent resistance to drugs received as retreatment, white bars, all other DAAs.

### Figure S8 (a) Timing of first line failiure (b) HCV VL at first-line failure (c, i-iii) Time to failure on first-line by fixed duration vs variable-duration randomisation

S8 (a) **Timing of first-line failure**

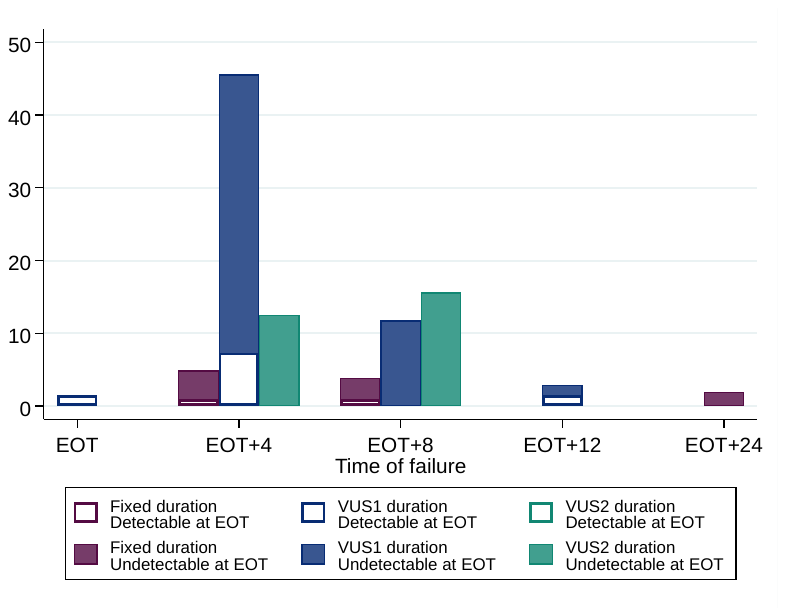

**(b)HCV VL at first-line failure**

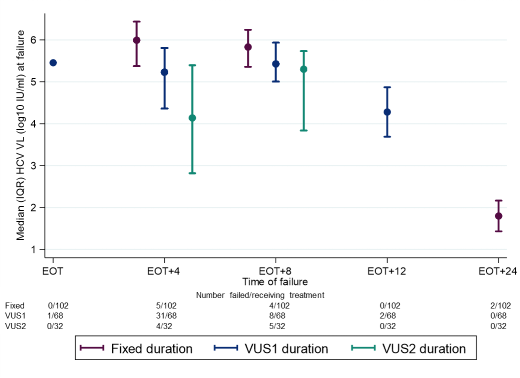

Note: HCV VL shown is the first, not the confirmatory, VL. No failures were considered re-infections after genome sequencing.

#### (C)(i) All failures

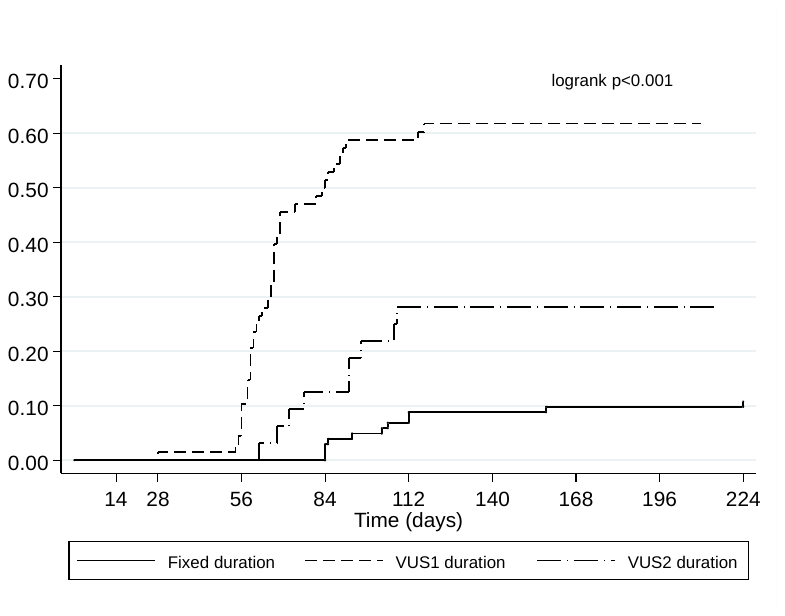

#### (C)(ii) Primary first line failure (never confirmed undetectable)

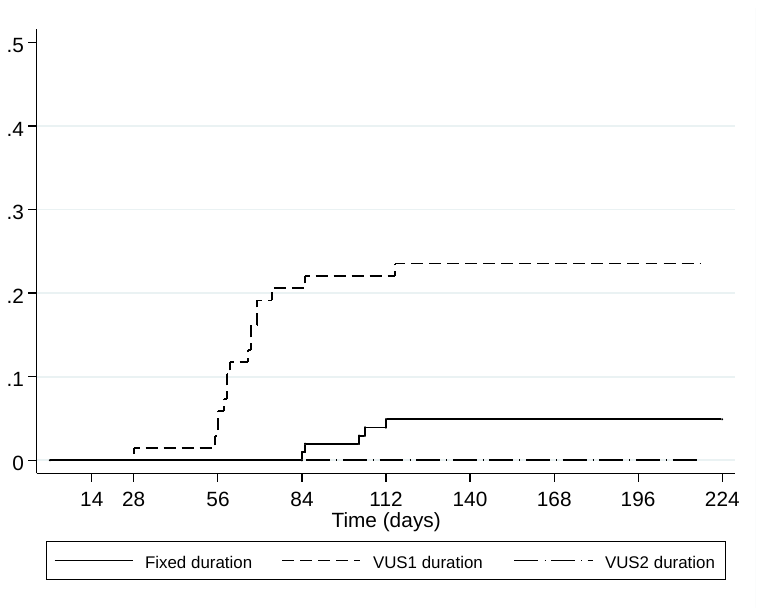

#### C(iii) HCV VL rebound (after confirmed undetectable)

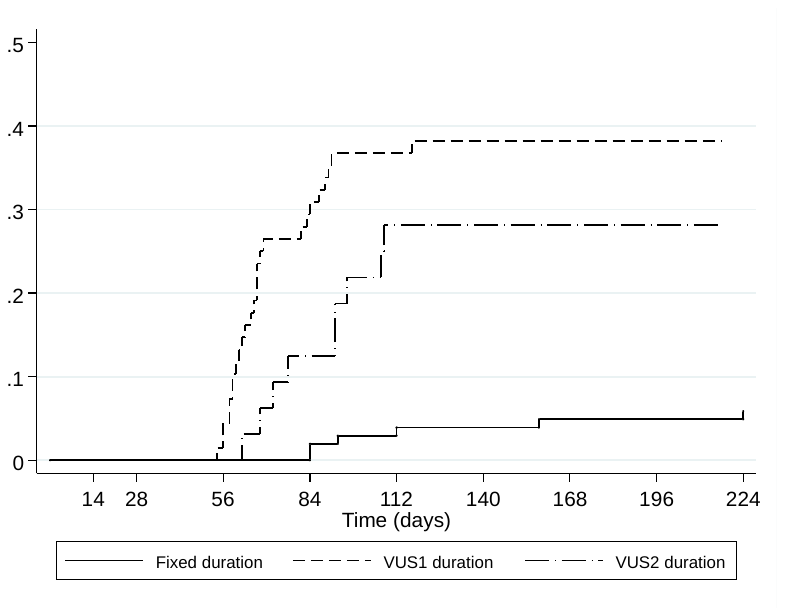

### Figure S9 Time to failure on first-line by ribavirin randomisation

#### (a) All failures

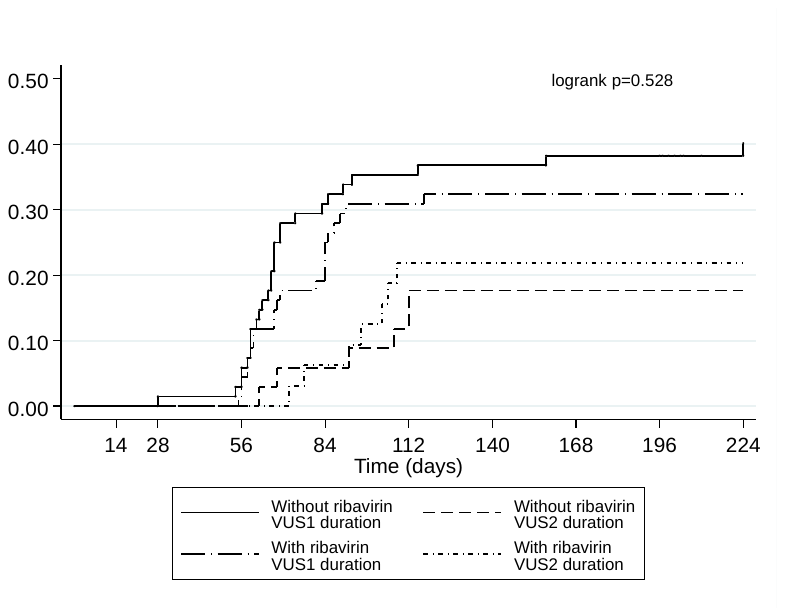

#### (b) Primary first-line failure (never confirmed undetectable)

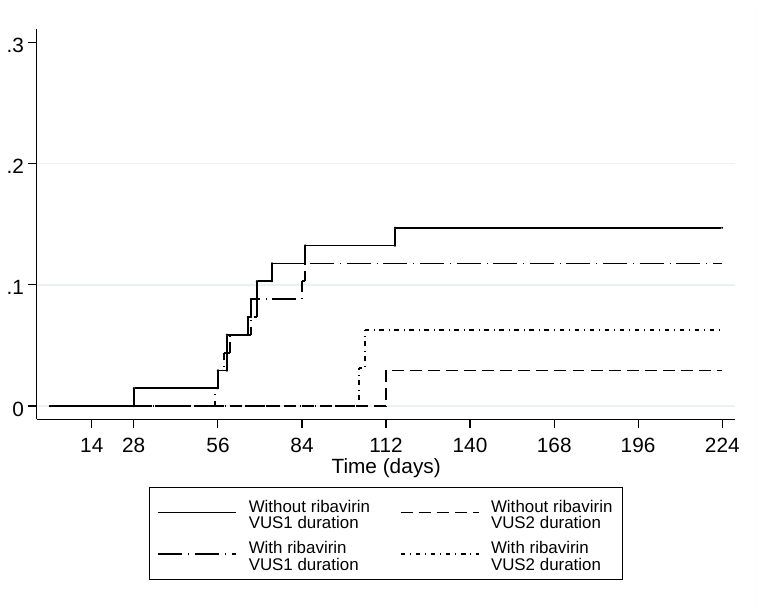

#### (c) HCV rebound (after confirmed undetectable)

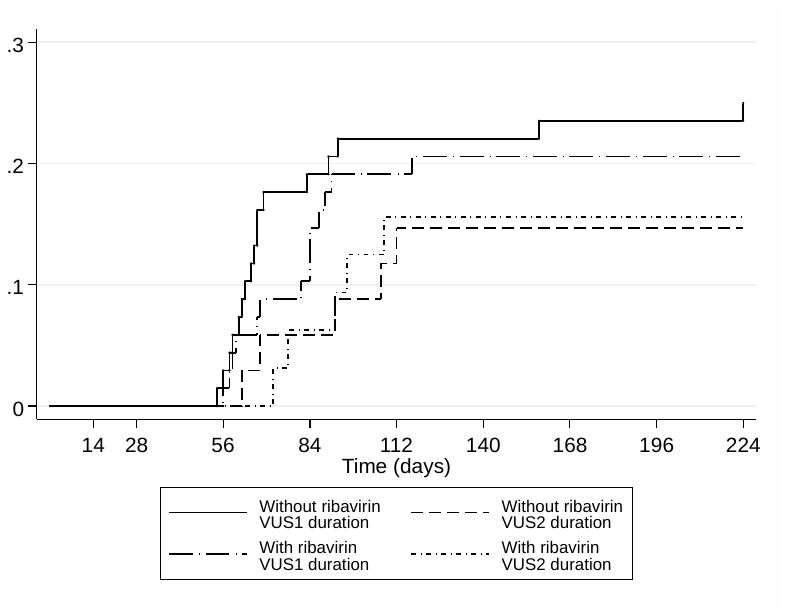

### Figure S10 HCV VL suppression on first-line by fixed-duration vs variable-duration randomisation

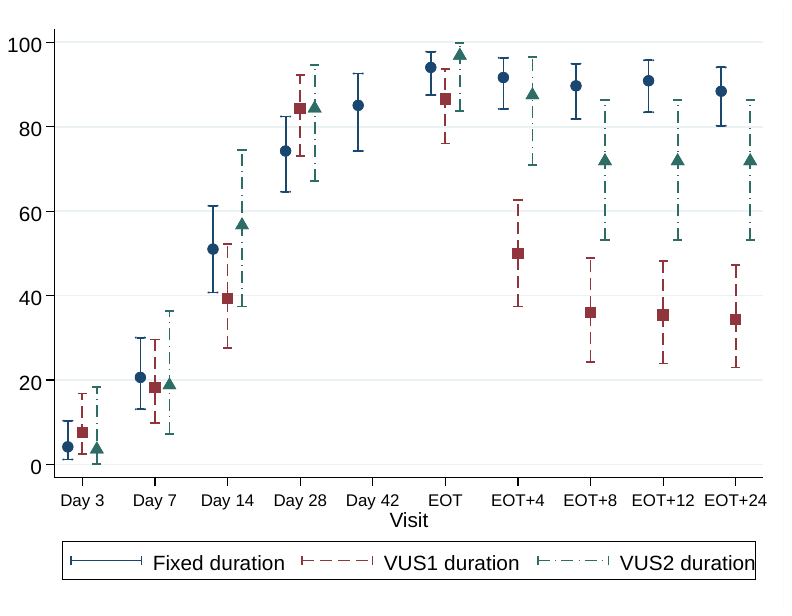

Note: carrying forward last detectable value for participants meeting failure criteria. No evidence of difference between groups through day 28 when all participants were receiving DAAs (global p=0.13 comparing fixed-duration vs variable-duration combined; p=0.10 and 0.82 comparing VUS1 and VUS2 vs fixed, respectively). Post-EOT global p<0.0001 comparing fixed-duration vs variable-duration combined; p<0.0001 and 0.53 comparing VUS1 and VUS2 vs fixed, respectively.

|  | Variable-duration (% undetectable) | Fixed-duration (% undetectable) | Total (% undetectable) | p-value |
| --- | --- | --- | --- | --- |
| Day 3 | 6/94 (6%) | 4/96 (4%) | 10/190 (5%) | p=0.53 |
| Day 7 | 18/98 (18%) | 20/97 (21%) | 38/195 (19%) | p=0.69 |
| Day 14 | 43/96 (45%) | 50/98 (51%) | 93/194 (48%) | p=0.38 |
| Day 28* | 81/96 (84%) | 75/101 (74%) | 156/197 (79%) | p=0.08 |
| Day 42 | - | 57/67 (85%) | 57/67 (85%) | - |
| EOT* | 89/99 (90%) | 95/101 (94%) | 184/200 (92%) | p=0.28 |
| EOT+4 weeks | 61/98 (62%) | 88/96 (92%) | 149/194 (77%) | p<0.001 |
| EOT+8 weeks | 46/96 (48%) | 87/97 (90%) | 133/193 (69%) | p<0.001 |
| EOT+12 weeks | 46/97 (47%) | 90/99 (91%) | 136/196 (69%) | p<0.001 |
| EOT+24 weeks | 45/96 (47%) | 84/95 (88%) | 129/191 (68%) | p<0.001 |

*For patients allocated 28-31 days, their EOT visit is also their 28 day visit. Included twice here.
NB: p-values calculated using chi-squared test or Fisher's exact test for small numbers.

### Figure S11 HCV VL suppression on first-line by ribavirin randomisation

#### (a) By ribavirin vs no ribavirin overall

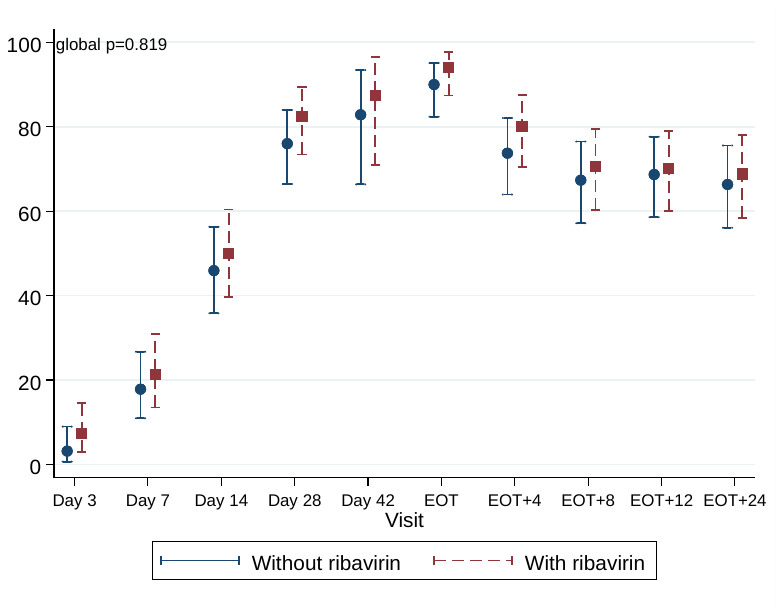

Note: carrying forward last detectable value for participants meeting failure criteria. No evidence of difference between groups through day 28 when all participants were receiving DAAs (global p=0.62 comparing ribavirin vs no ribavirin (combining fixed-duration vs variable-duration combined); interaction p=0.28). Post-EOT global p=0.48 comparing ribavirin vs no ribavirin (combining fixed-duration vs variable-duration combined; interaction p=0.22).

#### (b) For variable-duration patients only

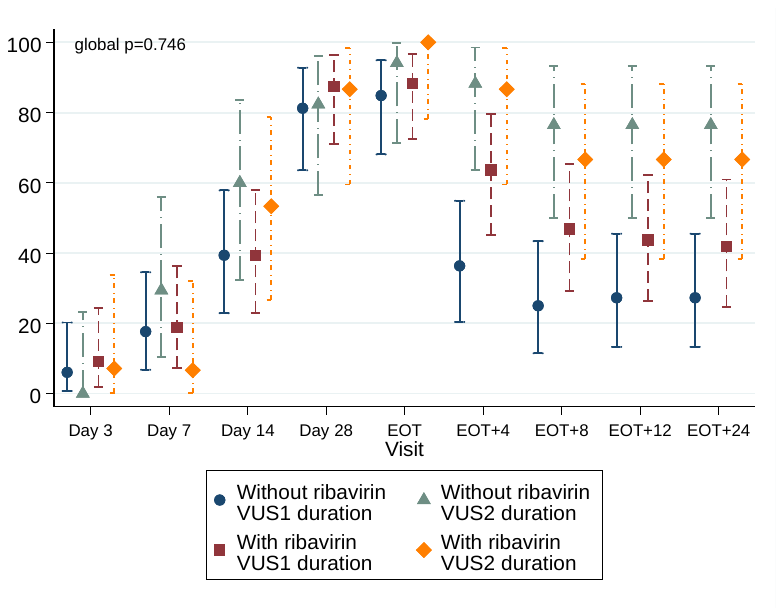

Note: carrying forward last detectable value for participants meeting failure criteria. No evidence of difference between groups through day 28 when all participants were receiving DAAs (global p=0.63 and 0.75 comparing ribavirin vs no ribavirin in VUS1 and VUS2, respectively). Post-EOT global p=0.28 and 0.83 comparing ribavirin vs no ribavirin in VUS1 and VUS2, respectively. No evidence of difference of ribavirin effect between VUS1 and VUS2 post-EOT (interaction p=0.17).

### Figure S12 Changes in laboratory test results

#### (a) Haemoglobin

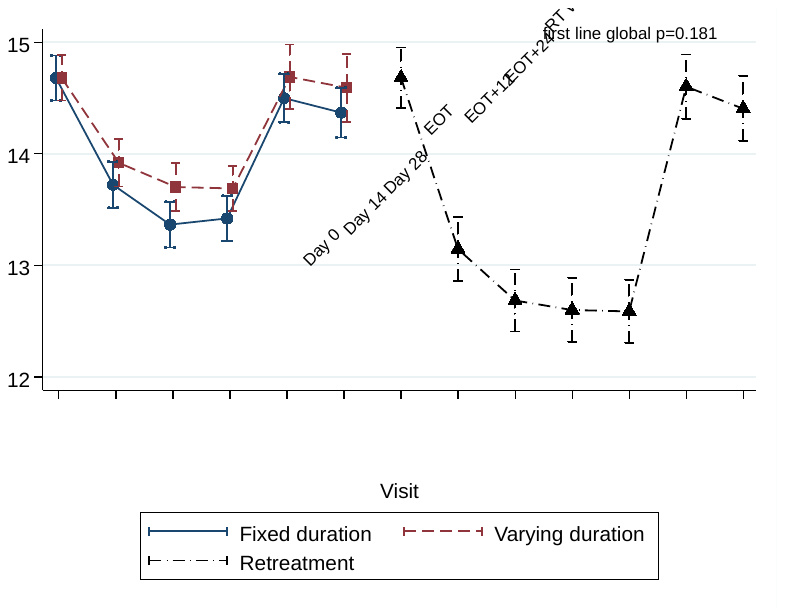

| First-line | Day 0 | Day 14 | Day 28 | EOT | EOT+12w | EOT+24w |
| --- | --- | --- | --- | --- | --- | --- |
| N: Variable | 100 | 92 | 91 | 99 | 50 | 45 |
| N: Fixed | 102 | 97 | 99 | 101 | 88 | 83 |
| p-value | - | 0.35 | 0.11 | 0.20 | 0.42 | 0.36 |

#### (b) ALT

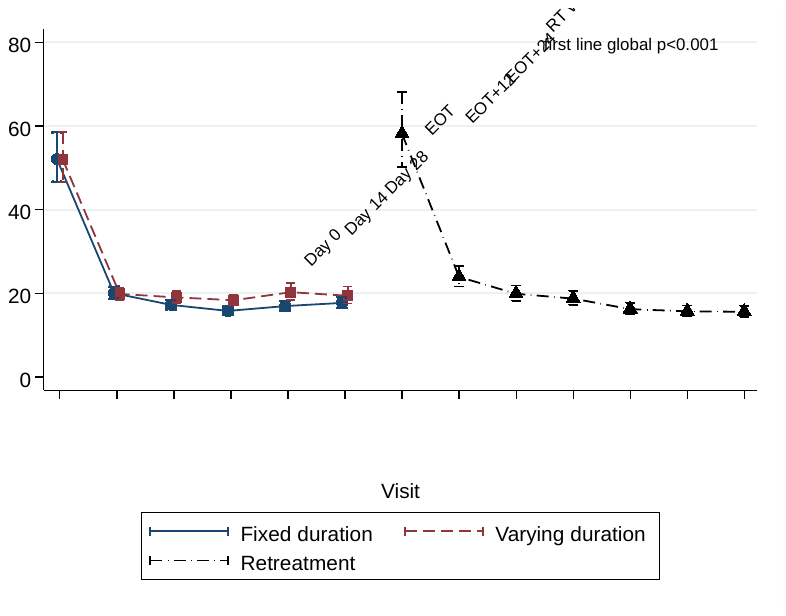

| First-line | Day 0 | Day 14 | Day 28 | EOT | EOT+12w | EOT+24w |
| --- | --- | --- | --- | --- | --- | --- |
| N: Variable | 100 | 92 | 91 | 98 | 51 | 45 |
| N: Fixed | 102 | 97 | 100 | 102 | 90 | 83 |
| p-value | - | 0.90 | 0.15 | 0.02 | 0.02 | 0.26 |

#### (c) AST

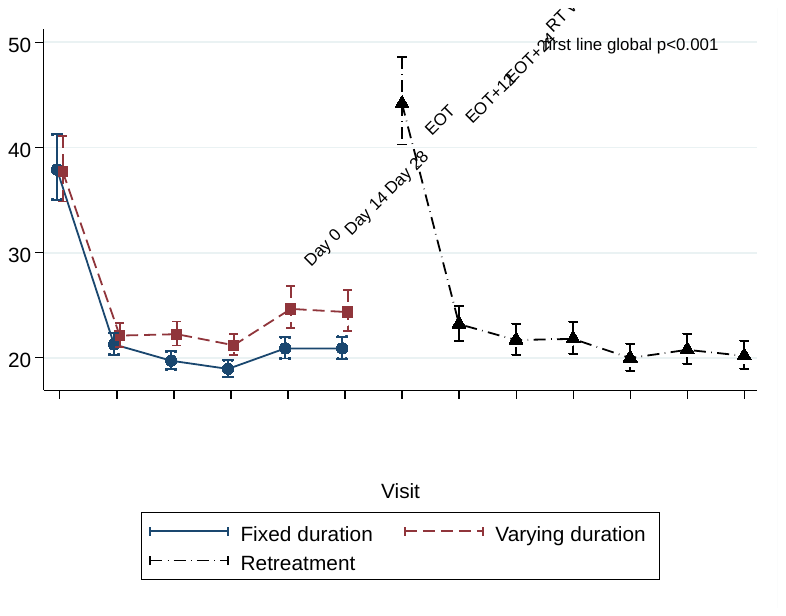

| First-line | Day 0 | Day 14 | Day 28 | EOT | EOT+12w | EOT+24w |
| --- | --- | --- | --- | --- | --- | --- |
| N: Variable | 90 | 77 | 79 | 87 | 40 | 40 |
| N: Fixed | 91 | 82 | 87 | 85 | 79 | 74 |
| p-value | - | 0.40 | 0.009 | 0.01 | 0.004 | 0.008 |

#### (d) Alkaline phosphatase

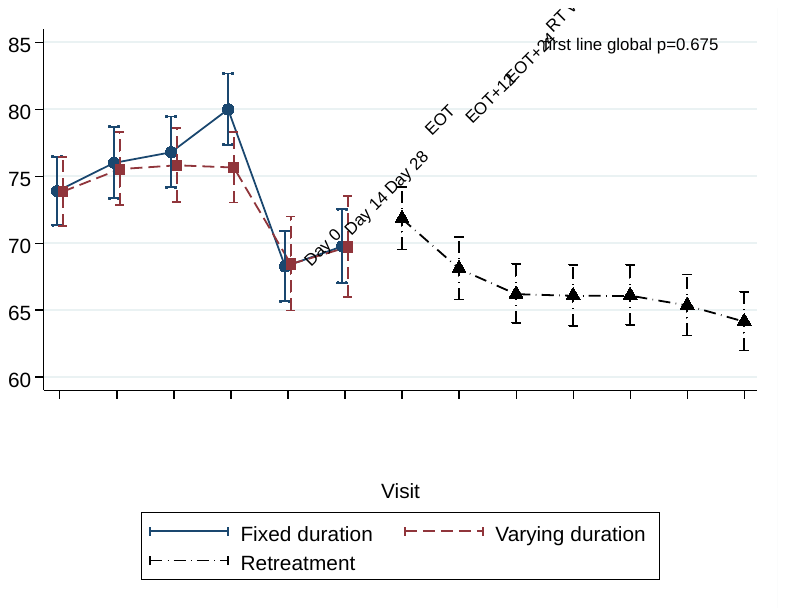

| First-line | Day 0 | Day 14 | Day 28 | EOT | EOT+12w | EOT+24w |
| --- | --- | --- | --- | --- | --- | --- |
| N: Variable | 100 | 92 | 91 | 98 | 51 | 45 |
| N: Fixed | 102 | 96 | 99 | 102 | 89 | 83 |
| p-value | - | 0.87 | 0.73 | 0.11 | 0.94 | 0.99 |

#### (e) Bilirubin

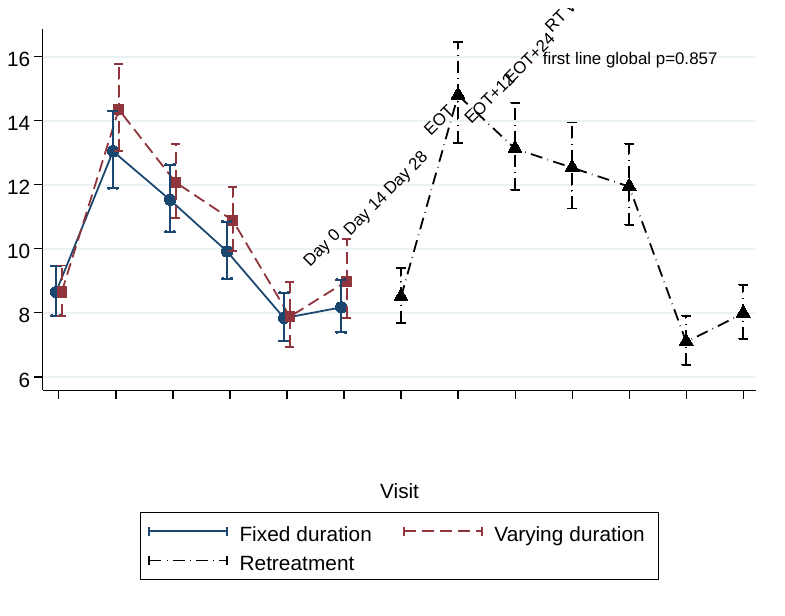

| First-line | Day 0 | Day 14 | Day 28 | EOT | EOT+12w | EOT+24w |
| --- | --- | --- | --- | --- | --- | --- |
| N: Variable | 100 | 92 | 91 | 97 | 51 | 45 |
| N: Fixed | 102 | 97 | 100 | 102 | 90 | 83 |
| p-value | - | 0.31 | 0.63 | 0.32 | 0.96 | 0.38 |

#### (f) eGFR

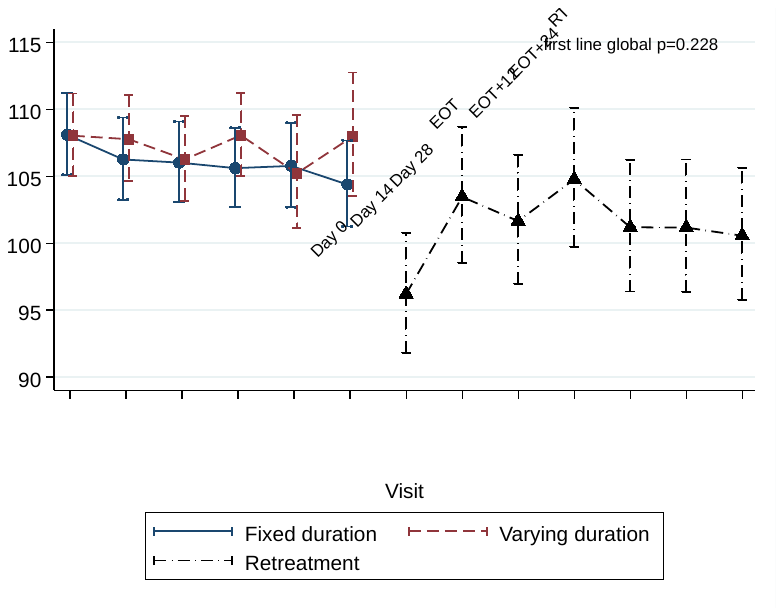

| First-line | Day 0 | Day 14 | Day 28 | EOT | EOT+12w | EOT+24w |
| --- | --- | --- | --- | --- | --- | --- |
| N: Variable | 100 | 92 | 91 | 99 | 50 | 45 |
| N: Fixed | 102 | 96 | 100 | 101 | 90 | 83 |
| p-value | - | 0.62 | 0.92 | 0.41 | 0.89 | 0.30 |

Note: For patients allocated 28-31 days, their EOT visit is also their 28 day visit and they are included twice in all analyses of change in laboratory values.

### Figure S13 Changes in laboratory test results by ribavirin randomisation

#### (a) Haemoglobin

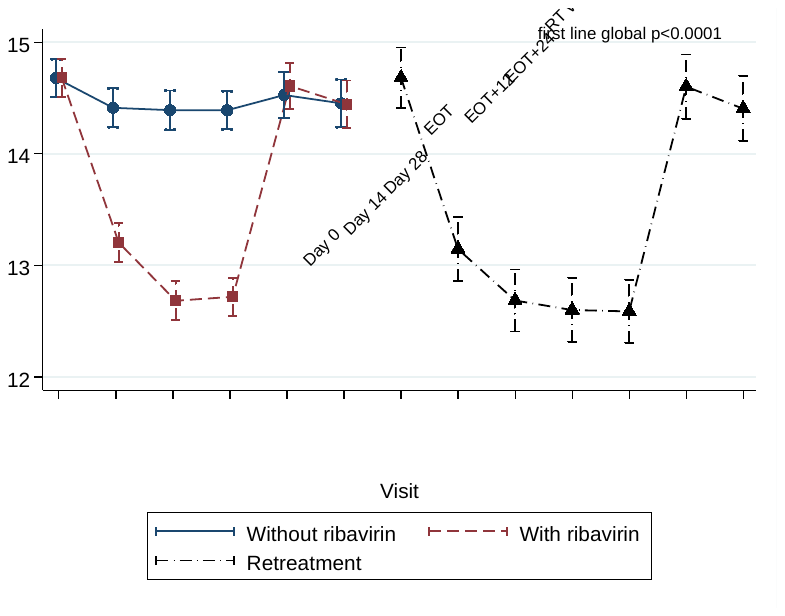

| First-line | Day 0 | Day 14 | Day 28 | EOT | EOT+12w | EOT+24w |
| --- | --- | --- | --- | --- | --- | --- |
| N: Variable | 100 | 93 | 96 | 100 | 70 | 64 |
| N: Fixed | 102 | 96 | 94 | 100 | 68 | 64 |
| p-value | - | <0.001 | <0.001 | <0.001 | 0.68 | 0.95 |

#### (b) ALT

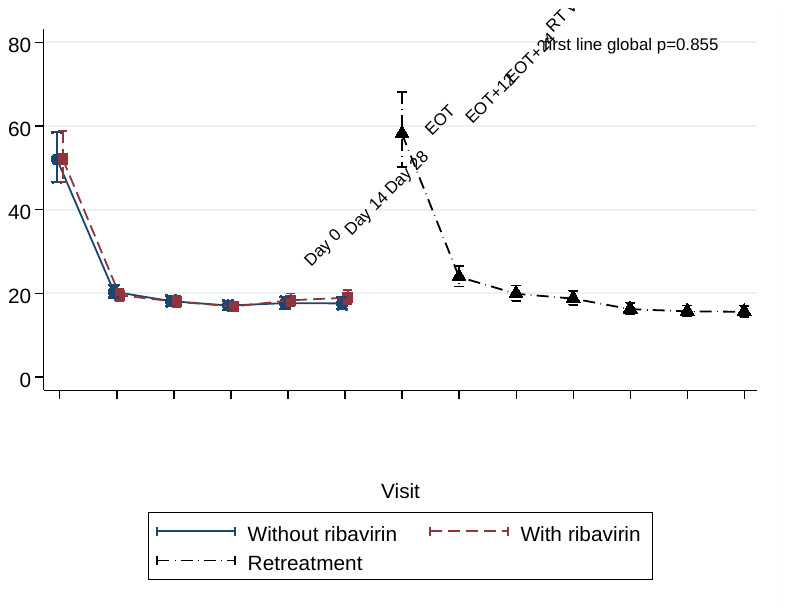

| First-line | Day 0 | Day 14 | Day 28 | EOT | EOT+12w | EOT+24w |
| --- | --- | --- | --- | --- | --- | --- |
| N: Variable | 100 | 93 | 96 | 99 | 71 | 63 |
| N: Fixed | 102 | 96 | 95 | 101 | 70 | 65 |
| p-value | - | 0.56 | 0.90 | 0.84 | 0.62 | 0.35 |

#### (c) AST

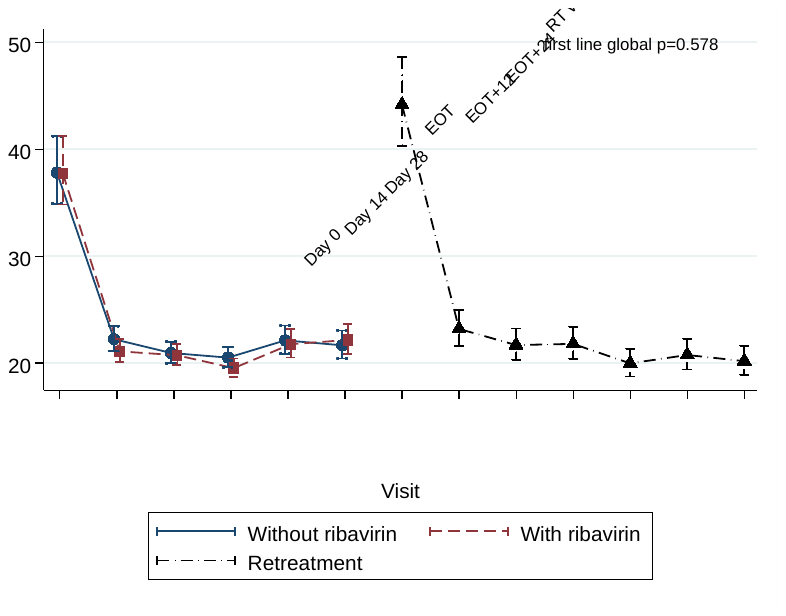

| First-line | Day 0 | Day 14 | Day 28 | EOT | EOT+12w | EOT+24w |
| --- | --- | --- | --- | --- | --- | --- |
| N: Variable | 89 | 78 | 82 | 86 | 57 | 57 |
| N: Fixed | 92 | 81 | 84 | 86 | 62 | 57 |
| p-value | - | 0.31 | 0.85 | 0.29 | 0.78 | 0.68 |

#### (d) Alkaline phosphatase

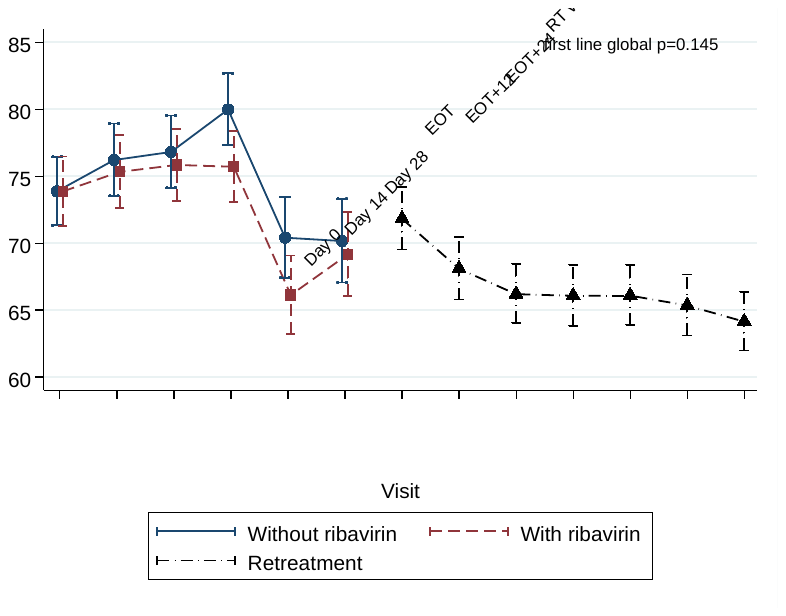

| First-line | Day 0 | Day 14 | Day 28 | EOT | EOT+12w | EOT+24w |
| --- | --- | --- | --- | --- | --- | --- |
| N: Variable | 100 | 93 | 95 | 99 | 70 | 63 |
| N: Fixed | 102 | 95 | 95 | 101 | 70 | 65 |
| p-value | - | 0.75 | 0.72 | 0.11 | 0.13 | 0.73 |

#### (e) Bilirubin

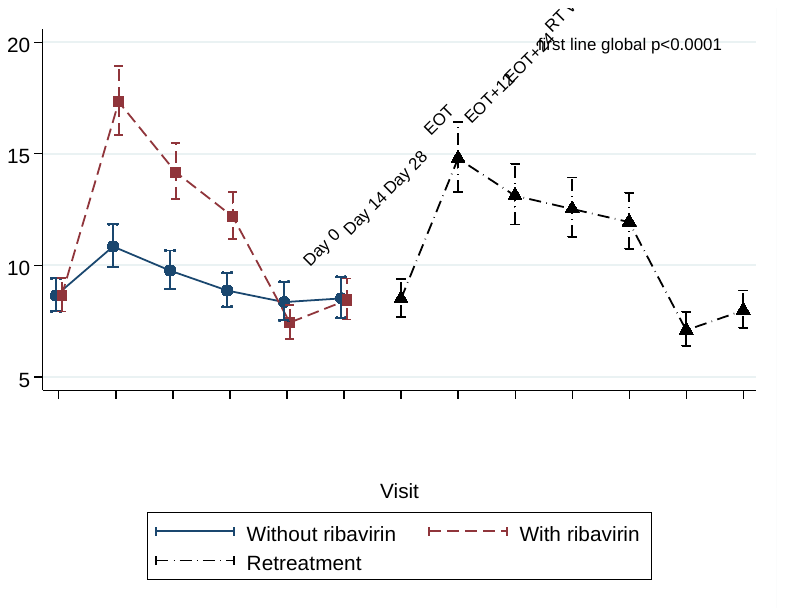

| First-line | Day 0 | Day 14 | Day 28 | EOT | EOT+12w | EOT+24w |
| --- | --- | --- | --- | --- | --- | --- |
| N: Variable | 100 | 93 | 96 | 98 | 71 | 63 |
| N: Fixed | 102 | 96 | 95 | 101 | 70 | 65 |
| p-value | - | <0.001 | <0.001 | <0.001 | 0.23 | 0.93 |

#### (f) eGFR

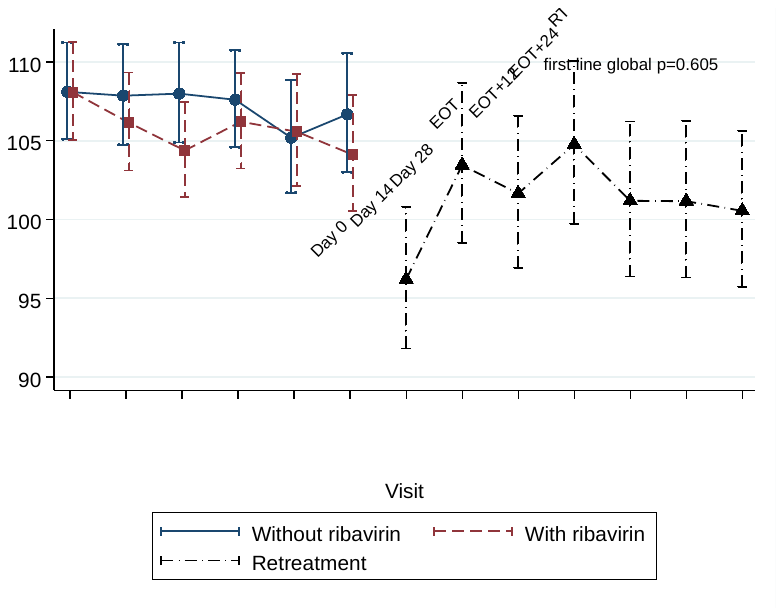

| First-line | Day 0 | Day 14 | Day 28 | EOT | EOT+12w | EOT+24w |
| --- | --- | --- | --- | --- | --- | --- |
| N: Variable | 100 | 94 | 96 | 99 | 71 | 63 |
| N: Fixed | 102 | 94 | 95 | 100 | 69 | 65 |
| p-value | - | 0.59 | 0.25 | 0.70 | 0.90 | 0.46 |

### Table S1 Duration of first-line treatment in the variable-duration group by protocol version

| From HCV VL (IU/ml) | To HCV VL (IU/ml) | Days if randomised before 01/04/2017 (VUS1) | Days if randomised after 01/04/2017 (VUS2) |
| --- | --- | --- | --- |
| LLOQ | 50,000 | 28 | 28 |
| 50,001 | 65,000 | 28 | 29 |
| 65,001 | 82,500 | 28 | 30 |
| 82,501 | 110,000 | 28 | 31 |
| 100,001 | 140,000 | 28 | 32 |
| 150,001 | 180,000 | 28 | 33 |
| 175,001 | 235,000 | 28 | 34 |
| 225,001 | 300,000 | 28 | 35 |
| 300,001 | 400,000 | 29 | 36 |
| 400,001 | 500,000 | 30 | 37 |
| 500,001 | 550,000 | 30 | 38 |
| 550,001 | 650,000 | 31 | 38 |
| 650,001 | 750,000 | 31 | 39 |
| 750,001 | 850,000 | 32 | 39 |
| 850,001 | 1,100,000 | 32 | 40 |
| 1,100,001 | 1,300,000 | 33 | 41 |
| 1,300,001 | 1,450,000 | 34 | 41 |
| 1,450,001 | 1,700,000 | 34 | 42 |
| 1,700,001 | 1,850,000 | 35 | 42 |
| 1,850,001 | 2,200,000 | 35 | 43 |
| 2,200,001 | 2,400,000 | 36 | 43 |
| 2,400,001 | 2,850,000 | 36 | 44 |
| 2,850,001 | 3,150,000 | 37 | 44 |
| 3,150,001 | 3,600,000 | 37 | 45 |
| 3,600,001 | 4,100,000 | 38 | 45 |
| 4,050,001 | 4,550,000 | 38 | 46 |
| 4,550,001 | 5,250,000 | 39 | 46 |
| 5,250,001 | 5,700,000 | 39 | 47 |
| 5,700,001 | 6,800,000 | 40 | 47 |
| 6,800,001 | 7,100,000 | 40 | 48 |
| 7,100,001 | 8,800,000 | 41 | 48 |
| 8,800,001 | upwards | 42 | 49 |

Note: VUS=variable ultra-short

### Table S2 Summary of length of treatment received in the variable duration arm

|  | VUS1 N=68 | VUS2  N=34 | Total N=102 |
| --- | --- | --- | --- |
| 4-5 weeks (28-34 days) | 48 (71%) | 11 (32%) | 59 (58%) |
| 5-6 weeks (35-42 days) | 18 (26%) | 13 (38%) | 31 (30%) |
| 6-7 weeks (42-49 days) | 2 (3%) | 10 (29%) | 12 (12%) |
| Mean (SD) | 32 (4.2) | 39 (5.6) | 35 (5.7) |

### Table S3 Additional characteristics at randomisation

|  | Total | VUS duration | Fixed-duration | Ribavirin | No ribavirin |
| --- | --- | --- | --- | --- | --- |
|  | N=202 | N=100 | N=102 | N=100 | N=102 |
| Weight (kg) | 74.0 (66.0, 84.6) | 73.0 (65.9, 84.1) | 76.1 (66.0, 85.9) | 69.9 (63.8, 82.3) | 78.9 (68.5, 86.8) |
| AST (IU/l) n=189 | 38 (30, 57) | 38 (29, 57) | 38 (31, 58) | 39 (31, 55) | 38 (29, 58) |
| ALP (IU/l) | 72 (59, 91) | 71 (59, 87) | 75 (59, 94) | 76 (61, 95) | 69 (58, 85) |
| eGFR (ml/min) | 109 (93, 131) | 109 (94, 126) | 109 (92, 138) | 107 (92, 126) | 110 (93, 133) |
| Total bilirubin (umol/l) | 9 ( 6, 12) | 8 (6, 11) | 9 (6, 12) | 9 (6, 12) | 9 (6, 12) |
| Previous HCV treatment |  |  |  |  |  |
| Ever spontaneously cleared and re-infected | 6 (3%) | 4 (4%) | 2 (2%) | 2 (2%) | 4 (4%) |
| Ever successfully treated with interferon and/or ribavirin and re-infected | 10 (5%) | 5 (5%) | 5 (5%) | 5 (5%) | 5 (5%) |
| Previously unsuccessfully treated with interferon and/or ribavirin | 24 (12%) | 12 (12%) | 12 (12%) | 11 (11%) | 13 (13%) |
| Intolerant relapser | 6 (26%) | 3 (25%) | 3 (27%) | 1 (10%) | 5 (38%) |
| Relapser after full treatment | 7 (30%) | 5 (42%) | 2 (18%) | 4 (40%) | 3 (23%) |
| Non-responder | 8 (35%) | 3 (25%) | 5 (45%) | 5 (50%) | 3 (23%) |
| Breakthrough on treatment | 2 (9%) | 1 (8%) | 1 (9%) | 0 | 2 (15%) |
| Modes of HCV infection |  |  |  |  |  |
| No known risk factor n=197 | 18 (9%) | 7 (7%) | 11 (11%) | 9 (9%) | 9 (9%) |
| Injecting drug use n=200 | 99 (50%) | 51 (51%) | 48 (48%) | 50 (51%) | 49 (49%) |
| Blood/blood products n=197 | 11 (6%) | 7 (7%) | 4 (4%) | 6 (6%) | 5 (5%) |
| Perinatal exposure n=197 | 4 (2%) | 0 | 4 (4%) | 3 (3%) | 1 (1%) |
| Known HCV positive sexual partner n=197 | 21 (11%) | 11 (11%) | 10 (10%) | 8 (8%) | 13 (13%) |
| Born abroad n=197 | 27 (14%) | 12 (12%) | 15 (15%) | 14 (14%) | 13 (13%) |
| High risk sexual partner n=201 | 71 (35%) | 37 (37%) | 34 (34%) | 35 (35%) | 36 (36%) |
| Tattoo n=197 | 27 (14%) | 14 (14%) | 19 (19%) | 14 (14%) | 19 (19%) |
| Healthcare exposure n=197 | 19 (10%) | 8 (8%) | 11 (11%) | 11 (11%) | 8 (8%) |
| Other n=196 | 20 (10%) | 11 (11%) | 9 (9%) | 10 (10%) | 10 (10%) |

Note: showing n (%) for categorical factors, or median (IQR) for continuous factors. Missing data indicated by denominators in the row label. As an indicator of imbalance, P>0.05 for all comparisons of baseline characteristics between groups other than weight (p=0.003) and ALP (p=0.04) between ribavirin and no ribavirin groups.

### Table S4 Characteristics at randomisation by VUS strategy under which randomisation occurred

|  | Randomised under VUS1† (N=136) | Randomised under VUS2 (N=66) | Total (N=202) |
| --- | --- | --- | --- |
| Randomised to VUS | 68 (50%) | 32 (48%) | 100 (50%) |
| Age (years) | 46.1 (38.1, 53.9) | 44.3 (36.8, 51.8) | 45.5 (37.5, 53.0) |
| Female at birth | 49 (36%) | 13 (20%) | 62 (31%) |
| Weight (kg) | 73.2 (64.6, 84.8) | 76.0 (68.0, 84.6) | 74.0 (66.0, 84.6) |
| BMI (kg/m^2^) | 25.0 (22.1, 27.6) | 24.4 (22.5, 27.0) | 24.9 (22.2, 27.2) |
| White ethnicity | 118 (87%) | 58 (88%) | 176 (87%) |
| Screening HCV viral load (IU/ml) | 755100 (244835,2344366) | 626862 (199526,1600000) | 711423  (218776,1995262) |
| Enrolment HCV viral load (IU/ml) n=199 | 770000 (251273,1872136) | 651594 (210000,1782738) | 741946 (249097,1872136) |
| HCV genotype/subgenotype |  |  |  |
| 1a | 108 (79%) | 58 (88%) | 166 (82%) |
| 1b | 28 (21%) | 6 (9%) | 34 (17%) |
| 4 | 0 | 2 (3%) | 2 (1%) |
| HIV co-infected | 38 (28%) | 30 (45%) | 68 (34%) |
| Fibroscan result (kPa) | 4.9 (4.1, 5.9) | 5.0 (4.4, 5.7) | 4.9 (4.2, 5.8) |
| Haemoglobin (g/dl) | 14.8 (14.2, 15.7) | 14.5 (13.6, 15.4) | 14.7 (14.0, 15.6) |
| ALT (IU/l) | 47 (33, 87) | 69 (42, 90) | 52 (34, 87) |
| AST (IU/l) n=189 | 38 (27, 58) | 40 (33, 54) | 38 (30, 57) |
| ALP (IU/l) | 72 (60, 90) | 72 (58, 95) | 72 (59, 91) |
| eGFR (ml/min) | 109 (92, 129) | 109 (94, 133) | 109 (93, 131) |
| Total bilirubin (umol/l) | 9 ( 6, 13) [2, 37] | 8 ( 6, 10) [3, 34] | 9 ( 6, 12) |
| IL28b genotype* |  |  |  |
| CC | 43 (32%) | 17 (26%) | 60 (30%) |
| CT | 74 (54%) | 32 (48%) | 106 (52%) |
| TT | 14 (10%) | 13 (20%) | 27 (13%) |
| No result | 5 (4%) | 4 (6%) | 9 (4%) |
| Resistance associated substitution (RAS) to any prescribed first-line drug n=188 | 20 (15%) | 7 (12%) | 27 (14%) |
| Previous HCV treatment |  |  |  |
| Ever spontaneously cleared and re-infected | 3 (2%) | 3 (5%) | 6 (3%) |
| Ever successfully treated with interferon and/or ribavirin and re-infected | 8 (6%) | 2 (3%) | 10 (5%) |
| Previously unsuccessfully treated with interferon and/or ribavirin | 21 (15%) | 3 (5%) | 24 (12%) |
| Intolerant relapser | 6 (29%) | 0 | 6 (26%) |
| Relapser after full treatment | 7 (33%) | 0 | 7 (30%) |
| Non-responder | 6 (29%) | 2 (100%) | 8 (35%) |
| Breakthrough on treatment | 2 (10%) | 0 | 2 (9%) |
| Modes of HCV infection |  |  |  |
| No known risk factor n=197 | 16 (12%) | 2 (3%) | 18 (9%) |
| Injecting drug use n=200 | 69 (51%) | 30 (45%) | 99 (50%) |
| Blood/blood products n=197 | 8 (6%) | 3 (5%) | 11 (6%) |
| Perinatal exposure n=197 | 4 (3%) | 0 | 4 (2%) |
| Known HCV positive sexual partner n=197 | 14 (11%) | 7 (11%) | 21 (11%) |
| Born abroad n=197 | 22 (17%) | 5 (8%) | 27 (14%) |
| High risk sexual partner n=201 | 43 (32%) | 28 (42%) | 71 (35%) |
| Tattoo n=197 | 22 (17%) | 5 (8%) | 27 (14%) |
| Healthcare exposure n=197 | 15 (11%) | 4 (6%) | 19 (10%) |
| Other n=196 | 9 (7%) | 11 (17%) | 20 (10%) |
| Current/recent alcoholism/alcohol abuse | 9 (7%) | 4 (6%) | 13 (6%) |
| Current/recent Illicit substance abuse | 38 (28%) | 26 (39%) | 64 (32%) |
| Treated with paritaprevir\ombitasvir\dasabuvir | 136 (100%) | 62 (94%) | 198 (98%) |
| Treated with paritaprevir\ombitasvir | 0 | 2 (3%) | 2 (1%) |
| Treated with glecaprevir\pibrentasvir | 0 | 2 (3%) | 2 (1%) |

† Randomised before 1 April 2017

* Result from whole genome sequencing or from Epistem POC test if genotyping result not available.

Note: showing n (%) for categorical factors, or median (IQR) for continuous factors. As an indicator of imbalance, P>0.05 for all comparisons of baseline characteristics between time periods of randomisation except for HCV genotype (excluding G4 (who could only be randomised into VUS2), p=0.049); sex, previous unsuccessful treatment with interferon and ribavirin (all p=0.02); and co-infected with HIV (p=0.01).

### Table S5 Summary of RAS to any DAA in genotype 1a by time point

|  | Baseline | Post-failure |
| --- | --- | --- |
| Resistance to NS5a inhibitors | N=13 | N=18 |
| 24: K24R | 1 (8%) | 2 (11%) |
| 28: M28T | - | 5 (28%) |
| M28V | 6 (46%) | 5 (28%) |
| 30: Q30H | 1 (8%) | - |
| Q30L | - | 1 (6%) |
| Q30R | - | 6 (33%) |
| 31: L31M | - | 2 (11%) |
| 58: H58D | - | 2 (11%) |
| 93: Y93F | 1 (8%) | - |
| Y93H | - | 1 (6%) |
| Y93N | 1 (8%) | 1 (6%) |
| Resistance to NS5b inhibitors | N=7 | N=3 |
| 448: Y448H | 2 (29%) | - |
| 556: S556G | 5 (71%) | 3 (100%) |
| Resistance to protease inhibitors | N=30 | N=14 |
| 36: V36M | 1 (3%) | 1 (7%) |
| 55: V55A | 3 (10%) | 2 (14%) |
| V55I | 3 (10%) | 1 (7%) |
| 80: Q80K | 21 (70%) | 9 (64%) |
| Q80L | 1 (3%) | - |
| 155: R155K | - | 1 (7%) |
| 168: D168A | - | 1 (7%) |

### Table S6 Summary of RAS to any DAA in genotype 1b by time point

|  | Baseline | Post failure |
| --- | --- | --- |
| Resistance to NS5a inhibitors | N=13 | N=2 |
| 30: R30Q | 1 (8%) | - |
| 31: L31I | 1 (8%) | - |
| L31M | 1 (8%) | - |
| 37: L37I | 1 (8%) | - |
| 54: Q54H | 11 (84%) | 2 (100%) |
| 58: P58S | 1 (8%) | - |
| Resistance to NS5b inhibitors | N=10 | N=3 |
| 159: L159F | 5 (50%) | 1 (33%) |
| 316: C316H | 2 (20%) | 1 (33%) |
| C316N | 4 (40%) | 1 (33%) |
| 556: S556G | 6 (60%) | 3 (100%) |
| Resistance to protease inhibitors | N=2 | N=1 |
| 122: S122N | 1 (50%) | 1 (100%) |
| 168: D168E | 1 (50%) | - |

### Table S7 Summary of adverse events by fixed-duration vs variable-duration randomisation

|  | Variable-duration | Fixed-duration | Total | p-value* |
| --- | --- | --- | --- | --- |
| Number randomised | N=100 | N=102 | N=202 |  |
| Median weeks follow-up (IQR) | 49.0  (29.0, 54.4) | 32.0  (32.0, 33.0) | 32.1  (31.2, 50.4) |  |
| **SAEs** | **5 (5%) [5]** | **5 (5%) [5]** | **10 (5%) [10]** | **p=1.00** |
| Life-threatening | 1 (1%) [1] | 1 (1%) [1] | 2 (1%) [2] |  |
| Required or prolonged hospitalisation | 5 (5%) [5] | 4 (4%) [4] | 9 (4%) [9] |  |
| Other important medical condition | 0 | 1 (1%) [1] | 1 (<1%) [1] |  |
| Relationship to trial drug (% of SAEs) |  |  |  |  |
| Unlikely | 2 (40%) | 2 (40%) | 4 (40%) |  |
| Not related | 3 (60%) | 3 (60%) | 6 (60%) |  |
| **Severe AEs** | **9 (9%) [16]** | **5 (5%) [5]** | **14 (7%) [21]** | **p=0.28** |
| Relationship to trial drug (% of severe AEs) |  |  |  |  |
| Definitely | 8 (50%) | 0 | 8 (38%) |  |
| Probably | 2 (13%) | 2 (40%) | 4 (19%) |  |
| Possibly | 3 (19%) | 0 | 3 (14%) |  |
| Unlikely | 0 | 1 (20%) | 1 (5%) |  |
| Not related | 3 (19%) | 2 (40%) | 5 (24%) |  |
| **AEs probably/definitely related to first-line drugs** | **3 (3%) [3]** | **1 (1%) [1]** | **4 (2%) [4]** | **p=0.37** |
| **AEs probably/definitely related to retreatment drugs** | **3 (3%) [7]** | **1 (1%) [1]** | **4 (2%) [8]** | **p=0.37** |
| **First-line drug changes due to AEs** | **3 (3%) [3]** | **1 (1%) [1]** | **4 (2%) [4]** | **p=0.37** |
| **Retreatment drug changes due to AEs** | **6 (6%) [11]** | **1 (1%) [1]** | **7 (3%) [12]** | **p=0.06** |
| **Grade 3/4 anaemia** | **3(3%) [3]** | **0** | **3 (1%) [3]** | **p=0.12** |

*p-values calculated using chi-square tests or Fishers exact test when numbers are small.

Note: no. of patients (% of patients) [no. of events]. Tables include data for both first-line and retreatment phases. For SAEs, HR=0.77 (0.21, 2.80) p=0.69. For severe (grade 3/4 AEs), HR=1.74 (0.58, 5.24) p=0.33.

### Table S8 Details of adverse events

| **Event** | **Variable, ribavirin** | **Variable, no ribavirin** | **Fixed, ribavirin** | **Fixed, no ribavirin** | **Total**  **Events** |
| --- | --- | --- | --- | --- | --- |
| **SAEs** |  |  |  |  |  |
| Accidental drug overdose* | 1 | 0 | 0 | 0 | 1 |
| Acute appendicitis | 0 | 0 | 0 | 1 | 1 |
| Adenocarcinoma in lower third of oesophagus* | 0 | 0 | 0 | 1 | 1 |
| Burn to foot - degree unknown | 1 | 0 | 0 | 0 | 1 |
| Liver abscess | 1 | 0 | 0 | 0 | 1 |
| Lower respiratory tract infection - pneumonia | 1 | 0 | 0 | 0 | 1 |
| Musculoskeletal pain in chest radiating to left arm | 0 | 0 | 0 | 1 | 1 |
| Pericarditis | 1 | 0 | 0 | 0 | 1 |
| R epididymo-orchitis | 0 | 0 | 0 | 1 | 1 |
| Urinary sepsis | 0 | 0 | 0 | 1 | 1 |
| **Severe AEs** |  |  |  |  |  |
| Abscess leg | 01 | 0 | 0 | 0 | 1 |
| Alcohol intoxication acute | 0 | 0 | 0 | 1 | 1 |
| Anaemia | 2 | 0 | 0 | 0 | 2 |
| Cellulitis of leg | 0 | 1 | 1 | 0 | 2 |
| Concentration loss | 1 | 0 | 0 | 0 | 1 |
| Haemoglobin low | 1 | 0 | 0 | 0 | 1 |
| Hyperbilirubinemia | 0 | 1 | 0 | 0 | 1 |
| Inguinal hernia | 1 | 0 | 0 | 0 | 1 |
| Insomnia | 3 | 0 | 0 | 0 | 3 |
| Jaundice | 0 | 0 | 0 | 1 | 1 |
| Lethargic | 1 | 0 | 0 | 0 | 1 |
| Low mood | 1 | 0 | 0 | 0 | 1 |
| Pyelonephritis | 0 | 0 | 1 | 0 | 1 |
| Sores mouth | 1 | 0 | 0 | 0 | 1 |
| Suicidal ideation | 1 | 0 | 0 | 0 | 1 |
| Syncope | 0 | 0 | 0 | 1 | 1 |
| Tinnitus | 1 | 0 | 0 | 0 | 1 |
| **AEs probably/definitely related to trial drugs** |  |  |  |  |  |
| Anaemia | 2 | 0 | 0 | 0 | 2 |
| Concentration loss | 1 | 0 | 0 | 0 | 1 |
| Haemoglobin low | 1 | 0 | 0 | 0 | 1 |
| Hyperbilirubinemia | 0 | 1 | 0 | 0 | 1 |
| Insomnia | 3 | 0 | 0 | 0 | 3 |
| Jaundice | 0 | 0 | 0 | 1 | 1 |
| Lethargic | 1 | 0 | 0 | 0 | 1 |
| Low mood | 1 | 0 | 0 | 0 | 1 |
| Syncope | 0 | 0 | 0 | 1 | 1 |
| Drug changes due to AEs | Variable-duration |  |  | Fixed-duration | Total |
| Anaemia | 3 | 3 | 0 | 0 | 6 |
| Concentration loss | 1 | 0 | 0 | 0 | 1 |
| Haemoglobin low | 1 | 0 | 1 | 0 | 2 |
| Hair loss | 0 | 0 | 1 | 0 | 1 |
| Hyperbilirubinemia | 0 | 1 | 0 | 0 | 1 |
| Insomnia | 1 | 0 | 0 | 0 | 1 |
| Lethargic | 1 | 0 | 0 | 0 | 1 |
| Low mood | 1 | 0 | 0 | 0 | 1 |
| Mouth ulcer | 1 | 0 | 0 | 0 | 1 |
| Sores mouth | 1 | 0 | 0 | 0 | 1 |

*Life-threatening events

### Table S9 Summary of adverse events by ribavirin randomisation

|  | With ribavirin | Without ribavirin | Total | p-value* |
| --- | --- | --- | --- | --- |
| Number randomised | N=100 | N=102 | N=202 |  |
| Median weeks follow-up (IQR) | 32.1  (30.4, 50.4) | 32.1  (31.9, 49.0) | 32.1  (31.2, 50.4) |  |
| **SAEs** | **5 (5%) [5]** | **5 (5%) [5]** | **10 (5%) [10]** | **p=1.00** |
| SAE criteria |  |  |  |  |
| Life-threatening | 1 (1%) [1] | 1 (1%) [1] | 2 (1%) [2] |  |
| Required or prolonged hospitalisation | 5 (5%) [5] | 4 (4%) [4] | 9 (4%) [9] |  |
| Other important medical condition | 0 | 1 (1%) [1] | 1 (<1%) [1] |  |
| Relationship to ribavirin (% of SAEs) |  |  |  |  |
| Unlikely | 2 (40%) | 2 (40%) | 4 (40%) |  |
| Not related | 3 (60%) | 3 (60%) | 6 (60%) |  |
| **Severe AEs** | **9 (9%) [15]** | **5 (5%) [6]** | **14 (7%) [21]** | **p=0.28** |
| Relationship to trial drug (% of severe AEs) |  |  |  |  |
| Definitely | 8 (53%) | 0 | 8 (38%) |  |
| Probably | 1 (7%) | 3 (50%) | 4 (19%) |  |
| Possibly | 3 (20%) | 0 | 3 (14%) |  |
| Unlikely | 1 (7%) | 0 | 1 (5%) |  |
| Not related | 2 (13%) | 3 (50%) | 5 (24%) |  |
| **AEs probably/definitely related to first line drugs** | **3 (3%) [3]** | **1 (1%) [1]** | **4 (2%) [4]** | **p=0.37** |
| **AEs probably/definitely related to retreatment drugs** | **2 (2%) [6]** | **2 (2%) [2]** | **4 (2%) [8]** | **p=1.00** |
| **First line drug changes due to AEs** | **4 (4%) [4]** | **0** | **4 (2%) [4]** | **p=0.06** |
| **Retreatment drug changes due to AEs** | **4 (4%) [8]** | **3 (3%) [4]** | **7 (3%) [12]** | **p=0.72** |
| **Grade 3/4 anaemia** | **3(3%) [3]** | **0** | **3 (1%) [3]** | **p=0.12** |

*p-values calculated using chi-square tests or Fishers exact test when numbers are small.

Note: no. of patients (% of patients) [no. of events]. Tables include data for both first-line and retreatment phases. For SAEs, HR=1.05 (95% CI 0.30, 3.63) p=0.94. For severe (grade 3/4 AEs), HR=1.92 (0.64, 5.72) p=0.59.
